# Conveying tactile object characteristics through customized intracortical microstimulation of the human somatosensory cortex

**DOI:** 10.1101/2024.03.08.24303392

**Authors:** C. Verbaarschot, V. Karapetyan, C.M. Greenspon, M. Boninger, S.J. Bensmaia, B. Sorger, R. A. Gaunt

## Abstract

Microstimulation of the somatosensory cortex can evoke tactile percepts in people with spinal cord injury, providing a means to restore touch. While location and intensity can be reliably conveyed, two issues that prevent creating more complex naturalistic sensations are a lack of methods to effectively scan the large stimulus parameter space and difficulties with assessing percept quality. Here, we addressed both challenges with an experimental paradigm that enabled three individuals with tetraplegia to control their stimulation parameters in a blinded fashion to create sensations for different virtual objects. Participants felt they could reliably create object-specific sensations and reported vivid object-appropriate characteristics. Despite substantial overlap in stimulus parameter selections across objects, both linear classifiers and participants could match stimulus profiles with their respective objects significantly above chance without any visual cues. We conclude that while visual information contributes to the experience of artificial touch, microstimulation in the somatosensory cortex itself can evoke intuitive percepts with a variety of tactile properties. This novel self-guided stimulation approach may be used to effectively characterize percepts from future stimulation paradigms.

---

The ultimate goal of prosthetics research is to create an artificial limb that effortlessly replaces lost function by seamlessly integrating the device with a person’s existing sensorimotor system. Tactile feedback is a key element to achieve this goal^1–5^, and a much-desired feature amongst prosthesis users^6,7^. Intracortical microstimulation (ICMS) of the somatosensory cortex can evoke localized sensations on a person’s paralyzed or insensate hand^8–10^ and is a promising way to provide this feedback. ICMS delivers tactile information directly to the brain, making it an attractive option for people with spinal cord injury or high-level amputation. How to best translate tactile object characteristics into stimulation patterns is non-trivial due to a limited understanding of the neural processing of touch, the complexity of the stimulation parameter space, and hardware restrictions that limit our ability to replicate natural neural responses using electrical stimuli^11,12^. So far, most studies focusing on the psychophysics of ICMS have manipulated one stimulation parameter at a time. The majority of these studies have been conducted in non-human primates who cannot verbalize their experienced sensations^13–15^. In the few human studies that exist, participants reported sensations like “buzzing”, “tingle”, and “pressure” in response to simple stimulation^8,10,16^. However, many combinations of stimulus amplitude, frequency and (multi-electrode) location(s) have yet to be explored, in part because the number of combinations grows exponentially as parameters are added. This drives a need for more efficient methods to explore the quality of ICMS-evoked sensations than the commonly applied stimulate-and-report task.

Here, we present an interface that three individuals with tetraplegia used to design their own sensations. Participants controlled stimulation presentation by “touching” an object displayed on a tablet (Fig. 1a), creating a more realistic experimental context compared to previous research. Moreover, through the self-paced scanning of a pre-selected parameter space (Fig. 1b), participants could adjust stimulation parameters without having to complete a sensory survey after each new parameter, providing an efficient and engaging experimental paradigm^8,10,17,18^. Participants controlled the stimulation amplitude, frequency, “biomimetic factor” and “drag”, while remaining blinded to the parameters themselves^19^ (Fig. 1b-c). Changes in amplitude and frequency have been shown to influence the perceived quality and intensity of the evoked sensations^8,10,15,20^. Further, most ICMS trains have commonly comprised fixed amplitudes and frequencies, or a stimulus amplitude that varies linearly based on an input variable^8,10,16^. However, neural responses in somatosensory cortex are dominated by a large burst of activity at the onset and offset of object contact, while sustained contact generates relatively low-level activity^15^. Using such transients (“biomimetic factor”) may evoke more intuitive, pleasant, or familiar sensations as it seeks to mimic patterns of natural brain activity. Lastly, to simulate the experience of the participants’ hand moving across an object, we stimulated three electrodes sequentially; each electrode evoked a sensation on an adjacent area on the hand. To create diffuse temporal transitions between consecutive electrodes, we created a “drag” parameter that controlled the degree of stimulation overlap between electrodes (Fig. 1c). With these methods, we investigated whether more complex stimulation trains could evoke naturalistic tactile object characteristics and identified specific stimulation parameters that best matched different object interactions.

**Figure 1.**
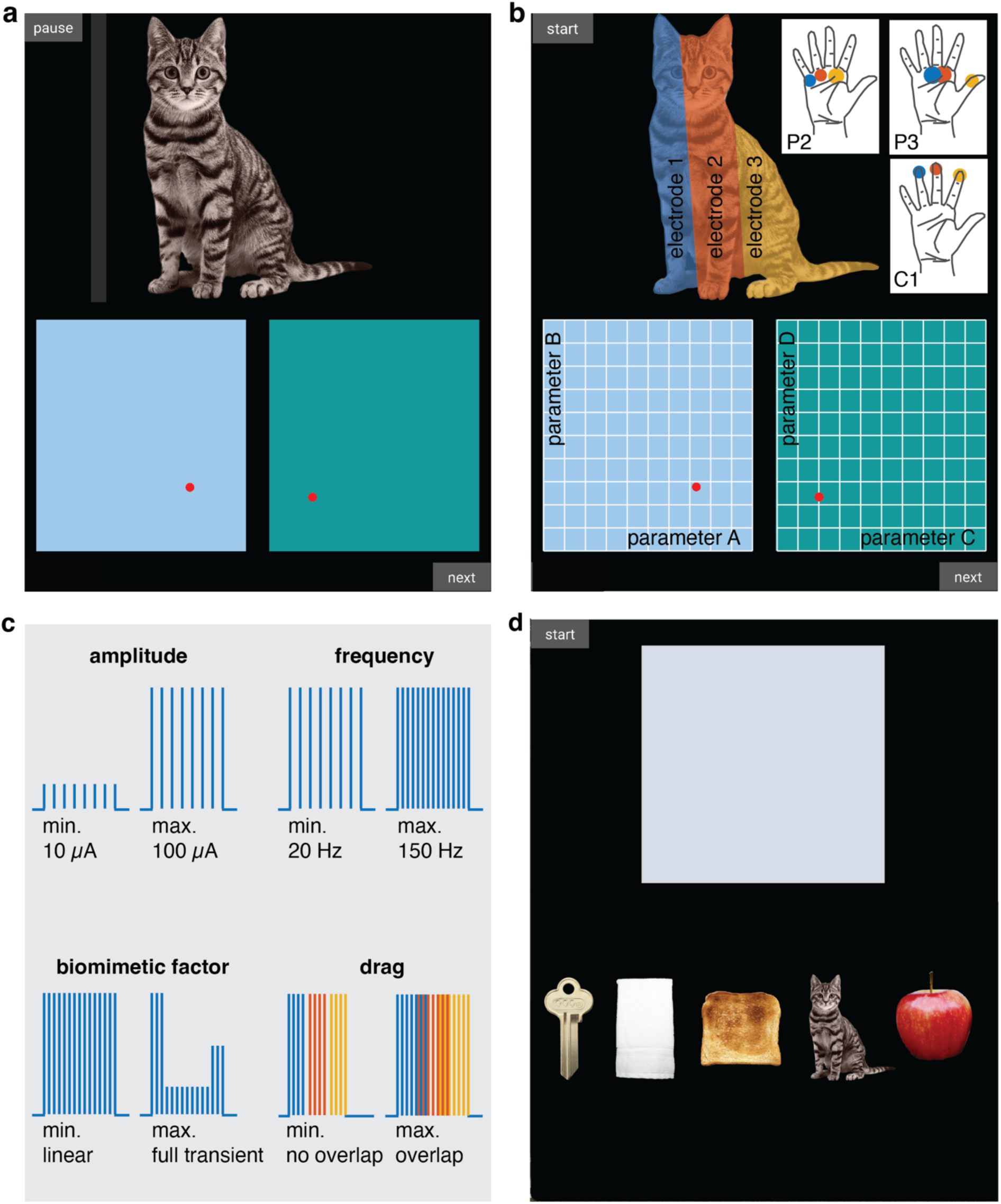
Tablet interface. **a.** Screenshot of the object-sensation mapping task. By changing the positions of the red cursors in the blue and green rectangles, the participant could adjust four stimulation parameters. Touching the object (‘cat’ in this example) generated stimulation trains in real-time. Participants manually touched the digital object or used an automated cursor by selecting the “start”/”pause” button in the top left corner, causing the grey bar to move across the object every 6.5 seconds. **b.** At the start of a trial, the four stimulation parameters (amplitude, frequency, biomimetic factor, drag) were randomly assigned to the A-D locations on the X- and Y-axes of the two rectangles. In addition, parameter values were randomly assigned to increase or decrease along the ten possible levels for each axis. Depending on where the participant touched the object, one of three stimulation electrodes was stimulated with the settings defined by the location of the cursors, evoking localized sensations on the palmar side of their right hand. The exact locations of the evoked sensations by each stimulation electrode were different for each participant, depending on the locations of the stimulation arrays in their somatosensory cortex. For more details on the exact implant locations for each participant, see Figure 1 in ref. ^28^. **c.** Schematic illustration of the four stimulation parameters at their minimum and maximum levels: amplitude, frequency, biomimetic factor, and drag. Each parameter controlled the stimulation of three individual electrodes and could be changed across 10 levels, i.e., there were 10 unique values per parameter. **d.** Replay task. When the participant touched the grey rectangle, a previously chosen set of stimulation parameters was replayed, evoking a sensation across the participants’ right hand. The participant was asked to indicate which object best matched the experienced sensation.

By putting participants in direct control of their own brain stimulation, we hoped to keep people engaged in exploring a large parameter space to develop personalized object-specific stimulus parameters. Our results show that participants felt they could reliably create stimulus-driven sensations that had object-appropriate characteristics for the visually presented objects. While the specific stimulus parameters that people chose for each object often overlapped, they were nevertheless able to identify the correct object when stimulation was delivered without any visual context significantly above chance. Further, the performance of the participants in this perceptual classification task was very similar to the performance of linear classifiers trained to identify objects based on the stimulation parameters alone. Given the large (and blinded) parameter space that was included in this study, it is promising that even in this challenging environment, participants were able to create distinct object sensations. We conclude that more complex stimuli, like those presented here, unlock a greater perceptual space that may allow people to distinguish artificially perceived objects with increased precision and intuition.

## Results

Three participants with tetraplegia (P2, P3, C1) designed their own artificial tactile sensations to represent interactions with a cat, apple, towel, piece of toast, and key. These objects were chosen because they spanned a range of tactile dimensions (compliance, temperature, friction, moisture, macro texture, micro texture, pleasantness, familiarity), as judged by 34 people with intact somatosensation (Fig. S1a) in an online survey (Fig. S1b). The participants used residual function in their left hand to interact with a tablet interface that generated “touch” sensations on the palmer surface of their right hand (Fig. 1a). Whenever participants touched a virtual object with their left hand, stimulation trains were delivered through microelectrode arrays implanted into Brodmann’s area 1 of the left hemisphere. Three electrodes were driven by contact with different regions of the object, such that the sensations ‘moved’ across their right hand as they moved across the object (Fig. 1b). In addition, participants used their left hand to control the position of cursors in two rectangular spaces on a tablet. Using these cursors, participants controlled the stimulus amplitude, frequency, biomimetic factor, and drag parameters (Fig. 1b-c). The parameters were randomly assigned to each of the four axes at the start of a trial, blinding participants to which axis controlled which parameter^19^.

During the “object-sensation mapping task”, participants repeatedly selected stimulation parameters that best represented each object. At the end of each trial, participants were asked to rate their satisfaction with the created sensation. To assess the discriminability of the created sensations, participants completed an additional “replay task” at the end of each session (Fig. 1d). During this task, stimulation trains that were created earlier were replayed without the corresponding object image. Participants were then asked to select the object that best matched the experienced sensation. In addition to these same day replay tests, two additional sessions used the replay task to test the discriminability and stability of a selection of sensations that were created across different days.

### Stimulation parameter selections were object specific

To validate whether the stimulus patterns generated using the interface could produce a variety of perceptual qualities, we first investigated how participants explored the stimulus parameter space during individual trials. Participants P2 and C1 broadly explored the space, but also identified “hotspots” in individual trials where they focused their attention (Fig. 2a). The presence of hotspots suggests that certain parameter combinations evoked more appropriate sensations for a given object than others (Fig. 2b). Participants P2 and C1 spent similar times exploring the parameter space (Fig. 2c,d) and received a similar amount of stimulation (Fig. 2e). In contrast to P2 and C1, participant P3 spent significantly less time exploring and receiving stimulation (Fig. 2c,d).

**Figure 2.**
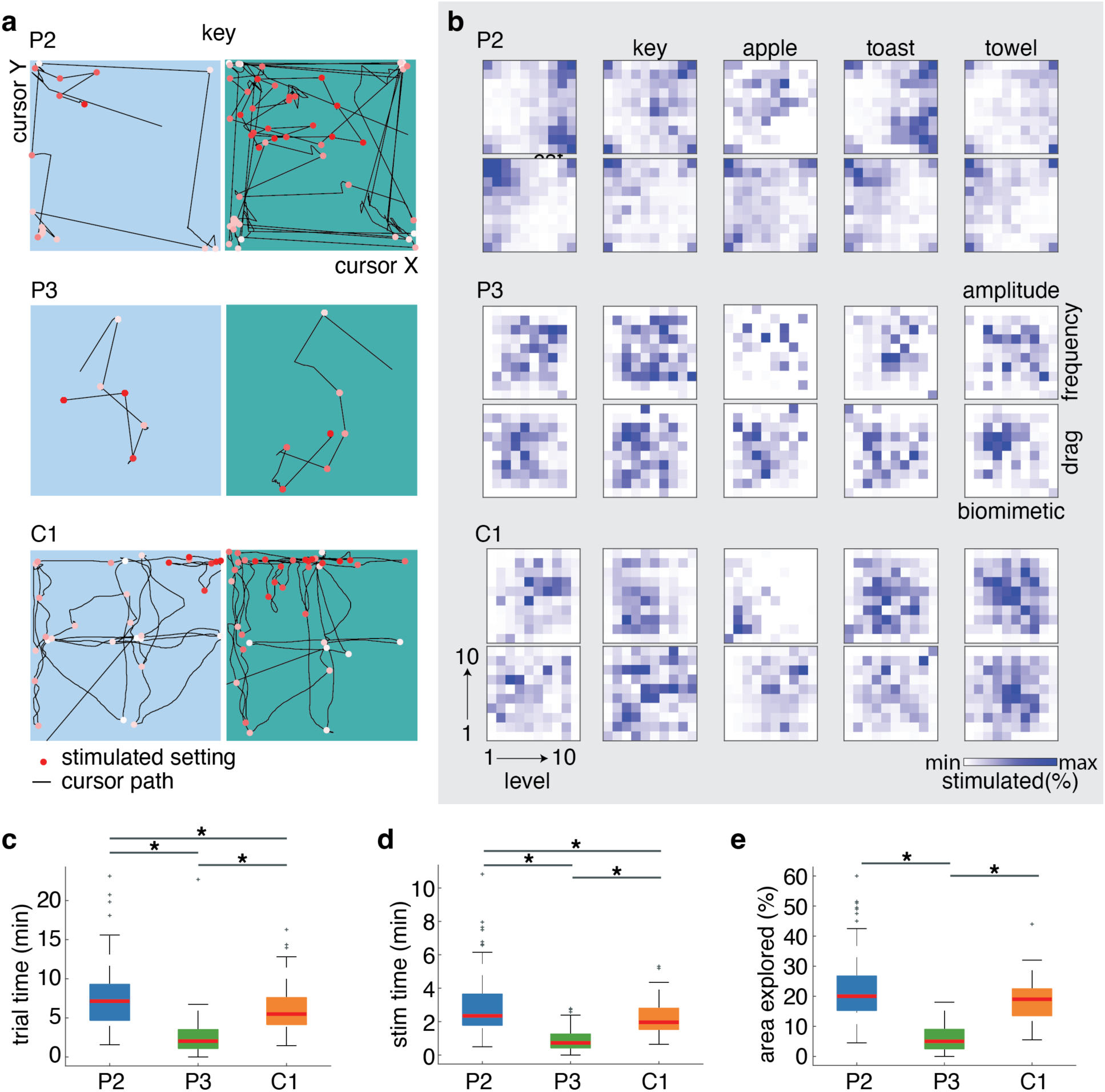
Exploration behavior of each participant using the interface. **a.** Participants P2, P3 and C1 behavior during an example object-sensation mapping trial for a key. Each dot indicates a stimulus parameter set during this single trial, changing in color from white (start trial) to red (end trial) as time progressed. The black lines show the exact movements of the cursor. P2 reported to explore the corners of each rectangle first followed by a more refined search starting from the most applicable corner; C1 reported to start from the middle of each rectangle and tested systematic changes in sensation as he moved away from the middle; P2 did not report a clear search strategy. Because the stimulation parameters were randomly assigned to the rectangle axes on each trial, the displayed cursor movements may refer to different parameters for each participant. **b.** Total percentage of time across all repetitions of each object spent on a certain stimulation setting for each participant. Parameter values increase from left (level 1) to right (level 10), and bottom (level 1) to top (level 10). Dark blue areas show “hotspots” within the parameter spaces where the participant spent most exploration time for a certain object. **c.** Trial completion times for each participant. At 6.0±2.5 minutes compared to 7.3±3.4 minutes, participant C1 was slightly faster than P2 (p = 0.001, Wilcoxon rank sum, Bonferroni corrected at α = 0.008, z = 3.22). At 2.4±2.4 minutes, P3 took significantly less time per trial compared to both P2 (p = 0.014^-27^, z = 11.30) and C1 (p = 0.026^-18^, z = -9.23). **d.** Total stimulation time within each trial for all participants. At 2.1±0.6 compared to 2.5±1.4 minutes, participant C1 had slightly less stimulation than P2 (p = 0.004, Wilcoxon rank sum, Bonferroni corrected at α = 0.008, z = 2.87). At 54±40 seconds, participant P3 had much less stimulation compared to both P2 (p = 0.044^-25^, z = 10.78) and C1 (p = 0.011^-18^, z = -9.32). **e.** Percentage of the parameter space explored per trial across all participants. At 19±7% compared to 22±11%, participant C1 explored a similar amount of the parameters space as P2 (p = 0.029, Wilcoxon rank sum, z = 2.18). At 6±5%, participant P3 explored much less than both P2 (p = 0.042^-32^, z = 12.18) and C1 (p = 0.051^-26^, z = - 10.97).

The variance in the participants’ parameter selections was high, leading to substantial parameter overlap between objects (Fig. 3). There were only a few instances where there were statistically significant differences in the distributions of stimulation parameters for different objects. For participant P2, there were significant differences across the objects in amplitude (p = 0.001^-1^, two-sided Kruskal-Wallis test Bonferroni corrected at α = 0.006, χ2(4) = 23.16, Fig. 3), biomimetic factor (p = 0.003, χ2(4) = 16.13, Fig. 3) and drag (p = 0.004, χ2(4) = 15.50, Fig. 3), while participant C1 showed significant differences in the distributions for amplitude only (p = 0.002^-1^, χ^2^(4) = 21.86, Fig. 3). For P3 no significant differences were found (Table S1).

**Figure 3.**
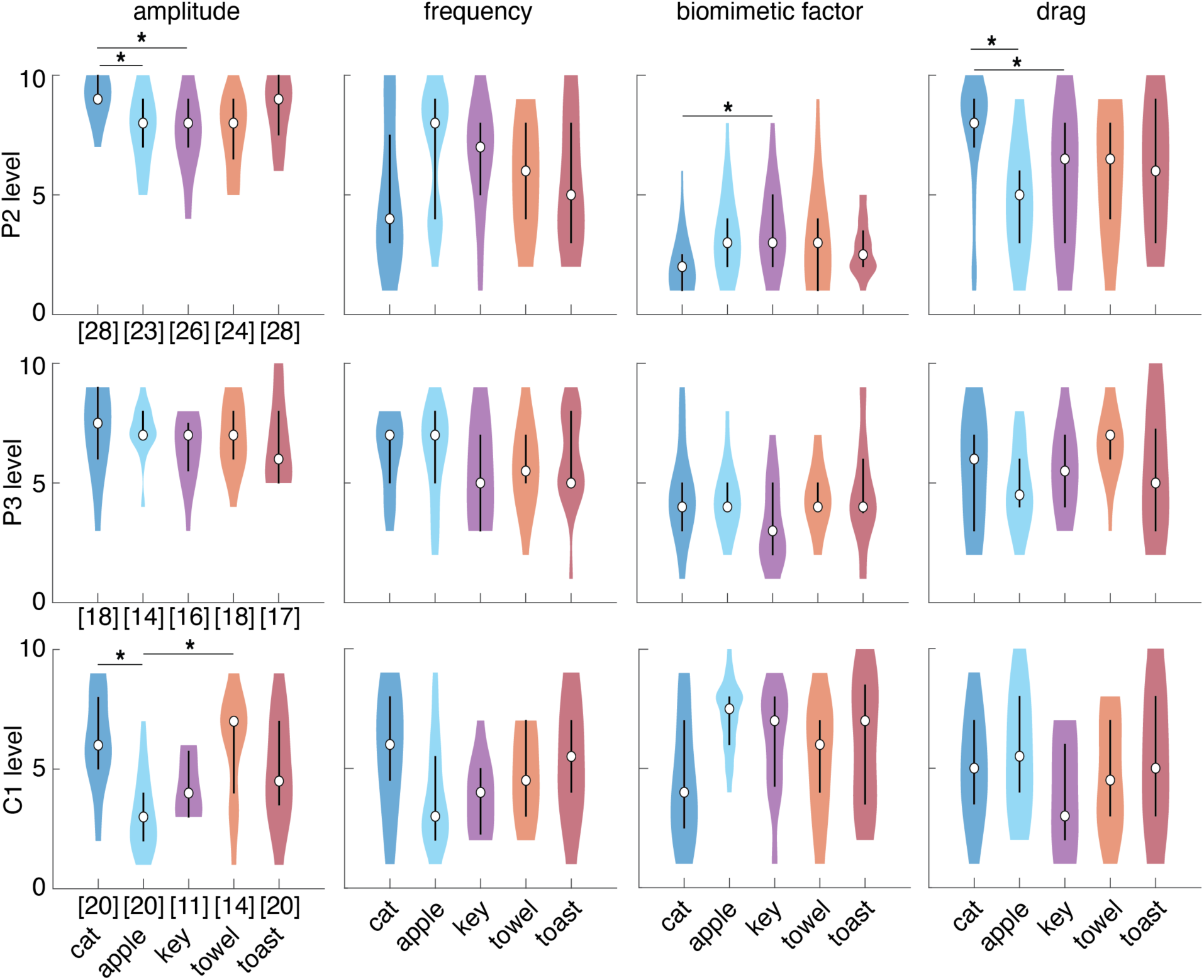
Stimulus parameter selections for each object, per participant. These data include only those trials that had a normalized satisfaction score of at least 50 out of 100, collected across 22 individual test days for participant P2 and 10 test days for participants P3 and C1. The number of repetitions per object is indicated in brackets for each object and participant. Each colored violin plot shows the distribution of a single parameter across all repetitions of that object. The median parameter value per object is displayed as an open black circle and the 25-75% quartiles are displayed as a thick black line. For P2, the amplitude was significantly different between a cat and apple (p = 0.004, one-sided Kolmogorov-Smirnov test Bonferroni corrected at α = 0.005, D = 0.48) as well as a cat and key (p = 0.008^-1^, D = 0.52). The biomimetic factor was significantly different for a cat and key (p = 0.002, D = 0.48), and the drag was significantly different for a cat and apple (p = 0.001^-1^, D = 0.59). For participant C1, the only difference was that the amplitude was significantly different for a cat and apple (p = 0.003^-1^, D = 0.64).

In contrast to the high overlap in individual stimulation parameters, the combination of stimulation parameters led to more separability amongst objects. To assess the distinctiveness of these combined parameter settings, we trained a linear discriminant analysis (LDA) classifier to identify which of the five objects a specific set of parameters belonged to using 10-fold cross validation. Across all five objects, the stimulation parameters predicted the object identity with an accuracy of 34±12% for participant P2 (p = 0.002, one-sided permutation test with 100 permutations without replacement, α = 0.05, Fig. 4a,c) and 37±15% for C1 (p = 0.002, Fig. 4a,e). At an accuracy of 21±13%, the stimulation parameters chosen by P3 predicted the associated object no better than chance (p = 0.341, Fig. 4a,d). These classification performances were not dominated by a single parameter. Rather, a combination of multiple parameters was required to achieve the maximum classification accuracy (Fig. 4b). Moreover, classification performance could not be explained simply by differences in the total charge delivered per electrode for each object (Fig. S2).

**Figure 4.**
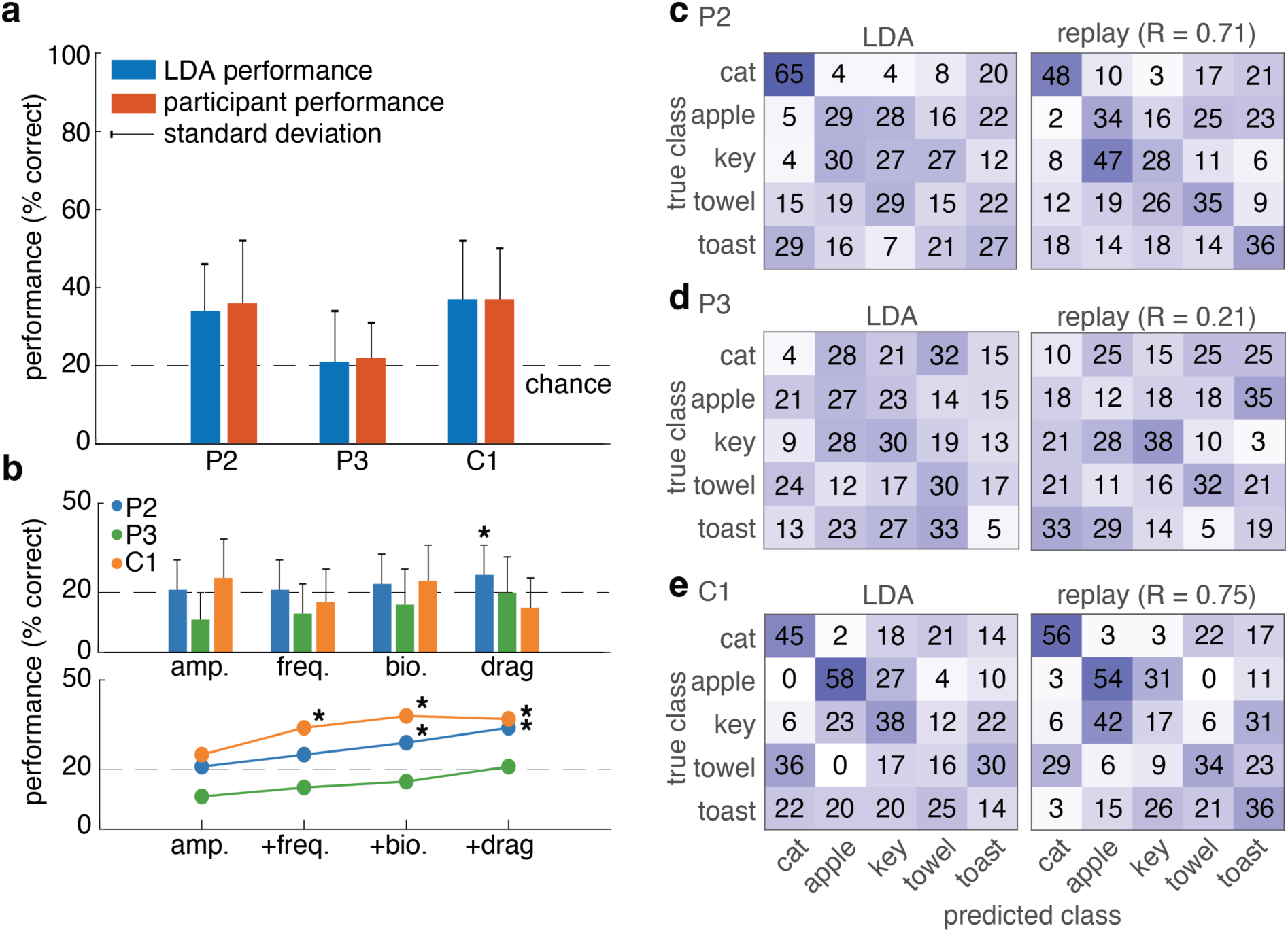
Effect of stimulation parameters on object LDA classification and participant replay performance. **a.** Participant and LDA performance on correctly identifying which of five objects was represented by a set of stimulus parameters. Vertical black lines indicate the standard deviation across bootstrapped LDA folds or replay sessions. **b.** LDA performance on individual (top) or different combinations (bottom) of stimulus parameters. The parameter combinations include: amplitude alone (leftmost); amplitude and frequency; amplitude, frequency and biomimetic factor; or all parameters (rightmost). Except for the P2’s drag parameter (p = 0.005, permutation test with 1000 permutations without replacement, Bonferroni corrected at α = 0.007, LDA accuracy = 25%), LDA predictions based on individual stimulation parameters were not significantly better than chance. Combinations of stimulation parameters enabled significant classification performances. For C1, LDA predictions based on amplitude and frequency were significant (p = 0.001, LDA accuracy = 34%). In addition, both P2 (p = 0.004, LDA accuracy = 29%) and C1 (p = 0.002, LDA accuracy = 38%) had significant LDA predictions based on a combination of amplitude, frequency and drag. **c.** Full confusion matrix for the LDA classifier and participant performance of P2. The R-value indicates the correlation between the participant and LDA performances. The numbers in each row indicate the percentage of times objects were correctly or incorrectly classified: for example, cat sensations that were classified by the LDA (left) or participant (right) as belonging to a cat, apple, key, towel or toast. **d.** Same as c., but for participant P3, **e.** Same as c., but for participant C1. There was a strong positive correlation of 0.71 and 0.75 between the perceptual replay and stimulus-parameter-based LDA performances of participant P2 and C1, and a low positive correlation of 0.21 for P3.

The object-specific stimulation parameters that drove LDA classifier performance also evoked distinct percepts that the participants could use to complete the replay task (Fig. 1d). Both participants P2 and C1 could use a given set of stimulus parameters to identify the corresponding object significantly above chance; P2 correctly identified the matching object with a mean accuracy of 36±16% (one-sided permutation test, 1000 permutations without replacement, α = 0.05, p = 0.001 Fig. 4a) per session and C1 did so with an accuracy of 37±13% (p = 0.001 Fig. 4a). As expected, participant P3 performed around chance with a mean accuracy of 22±9% (p = 0.051, Fig. 4a). For some objects, the participants created more distinctive stimulation parameter sets than others. This was reflected in the ability of LDA classifiers to accurately predict certain objects (Fig. 4c,d,e left column) as well as the ability of the participants to recall the correct object in the replay task (Fig. 4c,d,e right column). In fact, the perceptual performance of participants P2 and C1 of this within-session replay task closely matched those of the across-session stimulus-parameter-based LDA classifiers (Fig. 4a).

The significant performances of the LDA classifier for participants P2 and C1 suggest that there is some consistency in the selected object-specific parameters across sessions. To investigate the perceptual stability of the created sensations, we conducted two additional sessions of the replay task three (P2, C1) to seventeen (P3) weeks after finishing data collection. This time, we used three stimulus parameter sets per object that were created across previous sessions. All participants assigned two (P2, P3: cat, key) to three (C1: cat, key, towel) out 15 tested parameter sets consistently to the correct object. However, their overall replay performance did not rise above chance level (P2 - 22%, P3 - 19%, C1 - 25%). This suggests that although there was some consistency in the parameter sets of individual objects over time, their session-to-session variations influenced their perceptual experience. Indeed, the object-specific variations in stimulus parameters were found to be significantly larger across than within sessions (p = 0.002, one-sided Kruskal-Wallis test, α = 0.05, χ^2^(1) = 9.95, Fig. S3).

### Participants reported vivid and object-appropriate percepts

The participants described their sensory experiences at the end of each object-sensation mapping trial. These reports contained vivid object-appropriate sensory experiences. For example, P3 described the sensation for touching an apple as, “I found the perfect apple. It felt *light* but also *smooth*, *curved* and a little bit of *cool* and *wet*.”. Participant C1 described a cat sensation as, “very *light touch*, just *like petting a cat*. *Smooth silkiness* on fingertips. *Resistance* of cat. Has that *oily sensation*. It even has a sort of *warmth* to it.” Participant P2 described a towel sensation as, “It is *tingly*, but not quite *tappy*, but also not quite solid, reminds me of *roughness*.” Overall, participants described characteristic object qualities like “smooth”, “warm”, “dry” and “rough” in their sensation reports. These words are distinct from those used in previous reports of standard microstimulation trains^8,10,21^ and provide evidence of the experience of object-appropriate tactile characteristics in response to more complex customized microstimulation trains (Fig. 5).

**Figure 5.**
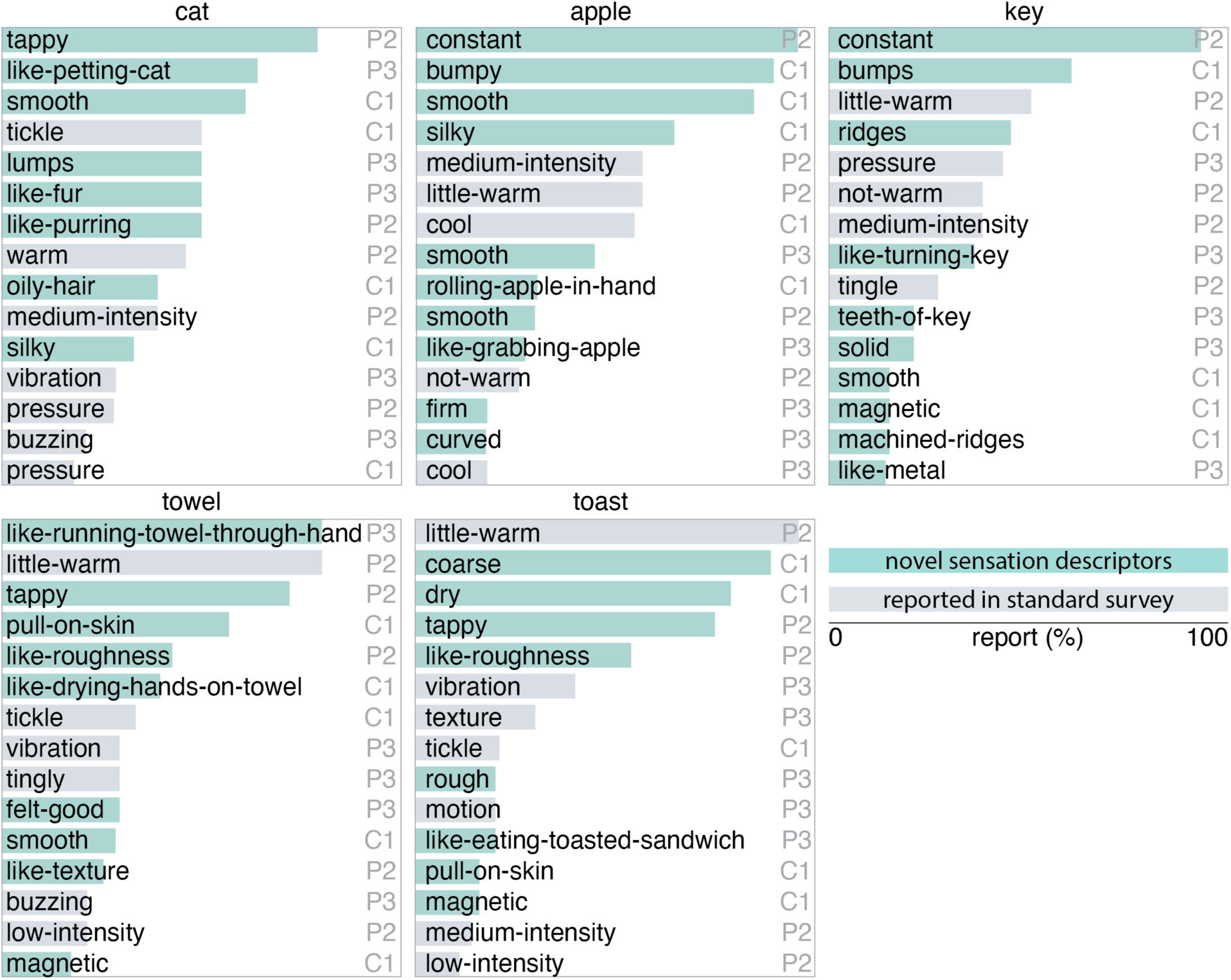
Verbal descriptors of the artificial sensations created using the interface. Each bar indicates the percentage of times participants used these descriptors in their report of a sensation for a specific object. Novel descriptors, compared to the those reported in previous ICMS studies that used standard stimulus trains that were devoid of a meaningful visual context^8,10,16^, are highlighted in green.

In addition, we asked participants to rate their satisfaction with the artificial sensations that they had created. Median normalized satisfaction scores were 68% for participant P2, 85% for P3 and 91% for C1 (Fig. 6a), indicating that participant P2 was the least satisfied and participant P3 the most satisfied with how well the ICMS-driven sensations represented the objects (Fig. 6a). There was a significant difference in the satisfaction scores between objects (p = 0.001, two-sided Kruskal-Wallis test, α = 0.025, χ^2^(4) = 18.26, Fig. 6b), which was driven by a difference in cats and keys (p = 0.005^-1^, two-sided Kolmogorov-Smirnov test Bonferroni corrected at α = 0.005, D = 0.33, Fig. 6b). No differences were found between the other object sensations.

**Figure 6.**
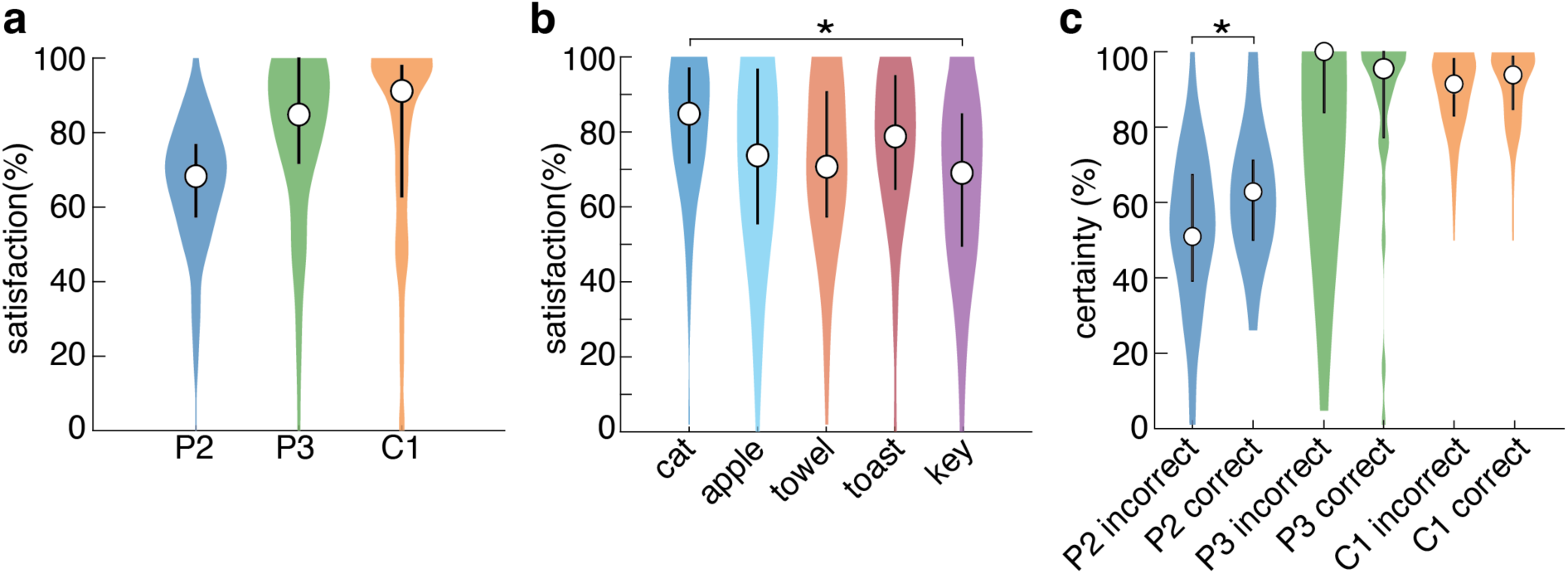
Normalized satisfaction and certainty ratings. **a.** Distribution of normalized satisfaction ratings for each participant. The median satisfaction score for each participant is displayed as an open circle. The 25-75% quartiles of the data are displayed as a thick line around the median. **b.** Normalized satisfaction ratings per object across all participants. On average, participants were more satisfied with the sensations for a cat than for a key (p < .001). No other differences were significant. **c.** Self-rated certainty with which a participant could (correctly or incorrectly) identify the object corresponding to a set of stimulus parameters in the replay task. Participant P2 was significantly more certain of his answers when they were in fact correct, compared to when they were not (p < .001).

During the replay task, we asked participants how certain they were of their decision. Both P3 and C1 were very certain of their answers (92% - 100%), regardless of whether they were correct (Fig. 6c); there was no significant difference in certainty between correct and incorrect trials (P3, p = 0.861, Wilcoxon rank sum, α = 0.025, z = -1.07; C1, p = 0.114, z = 1.21). In contrast, with a median of 63% for correct and 51% for incorrect selections, P2 was significantly more certain of his answer when he was correct (p = 0.003, z = 2.73, Fig. 6c). These results suggest that even in the absence of a visual context, participants experience subjectively distinct sensations in response to their object-specific parameter selections.

Lastly, participants completed a short survey on their overall experience with the interface. Participants reported that they liked the graphical interface, felt interested, involved, and drawn into the experimental task, found it fun and rewarding to do, and did not find the task frustrating, demanding, or confusing (Fig. S4). These results, together with the significant performances on the replay task, demonstrate that the tablet interface was an effective means to create vivid and object-appropriate sensations.

### Artificial percepts reflect naturalistic tactile features

To assess whether the quality of our participants’ artificial sensations reflected the natural tactile characteristics of objects, we compared them to reports of object qualities provided by people with intact somatosensation. Using an online survey (Fig. S1b), 34 people rated the compliance, temperature, friction, moisture, microstructure, and macrostructure of the objects included in this study. The five objects in this study had a range of tactile properties that varied across multiple dimensions (Fig. 7a). We investigated whether the participants’ stimulation parameter selections correlated with these expected object-specific characteristics. To do so, individual LDA classifiers were trained to make predictions about whether a particular stimulus parameter set represented specific object qualities, such as compliance (hard vs. soft), temperature (warm vs. cold), and macro texture (edged vs. round). Based on the stimulus parameters chosen by both P2 and C1, we could significantly predict binary levels of both object compliance (P2: 63±12% accuracy, p = 0.010, permutation test with 1000 permutations without replacement, α = 0.025; C1: 71±14% accuracy, p = 0.002, Fig. 7b) and temperature (P2: 72±11%, p = 0.001 C1: 76±13%, p = 0.001 Fig. 7b). LDA classifier performance was not significant for the other tactile characteristics (Table S2). These results suggest that participants attempted to design intuitive sensations, as their parameter selections correlated with expected differences in object compliance and temperature.

**Figure 7.**
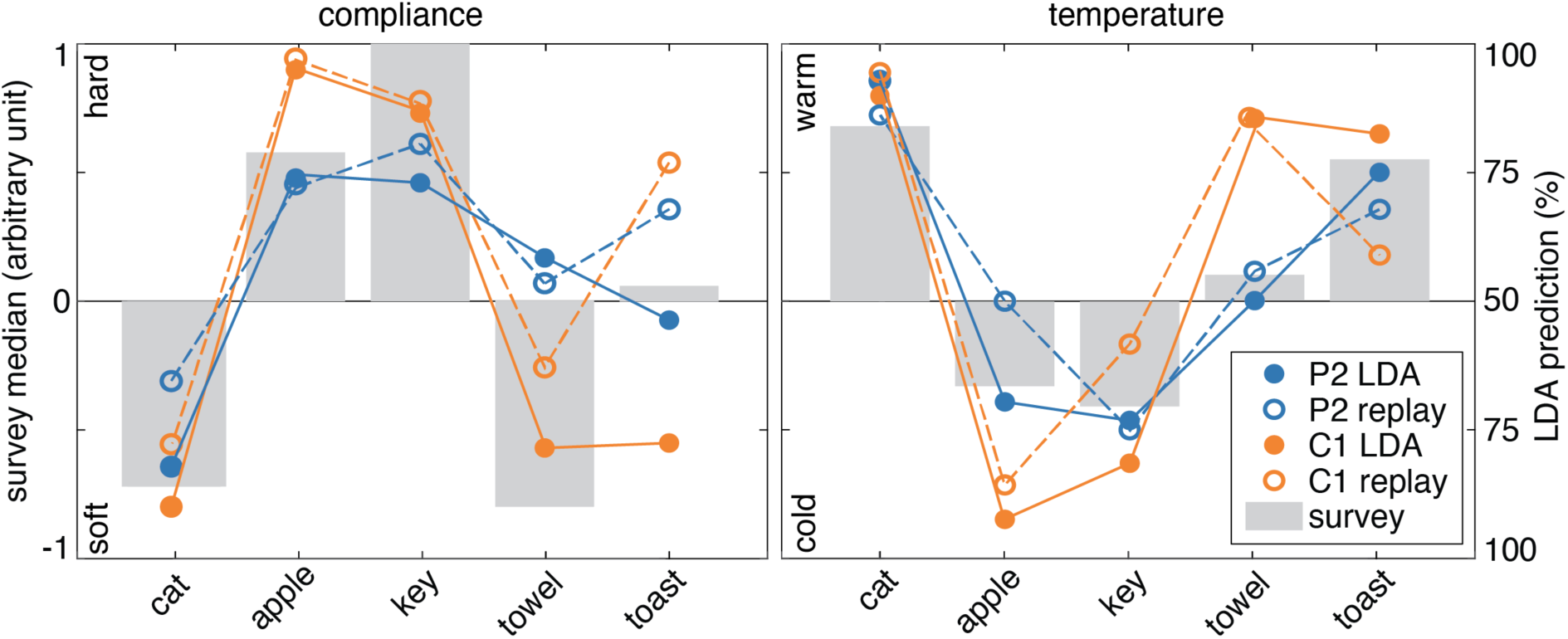
Expected versus predicted object characteristics. The gray bars show the median object quality survey ratings for compliance and temperature for each of the five objects. The filled circles indicate the percentage of stimulus parameter sets for each object that were ascribed a certain compliance (hard vs. soft) or temperature (warm vs. cold) by the LDA classifier for participant P2 and C1. For example, about 80-90% of all cat sensations were classified as being ‘soft’ whereas 75-95% of all apple sensations were classified as being ‘hard’. The open circles show how P2 and C1’s choices in the replay task matched the compliance and temperature predictions of the object quality survey. For example, C1 matched 78% of all their cat sensations with something soft (i.e. a cat or towel) in the replay task, whereas 97% of all apple sensations were matched with something hard (i.e. an apple, key or toast). Participant P3 is excluded from this follow-up analysis due to their chance level performance on the replay task.

The object quality survey results show that some objects used in the stimulation experiments had quality profiles that shared certain characteristics (Fig. 8a). Similarly, we found that during the replay task, participants were more likely to confuse some pairs of objects more than others (Fig. 8b). Moreover, sensations corresponding to the two most similar objects, as judged by the survey results, were significantly more likely to be confused by our participants in the replay task than the two most dissimilar objects (p = 0.004^-4^, two-sample Kolmogorov-Smirnov test, α = 0.025, D = 0.38, Fig. 8c), suggesting that participants tried to create intuitive object sensations.

**Figure 8.**
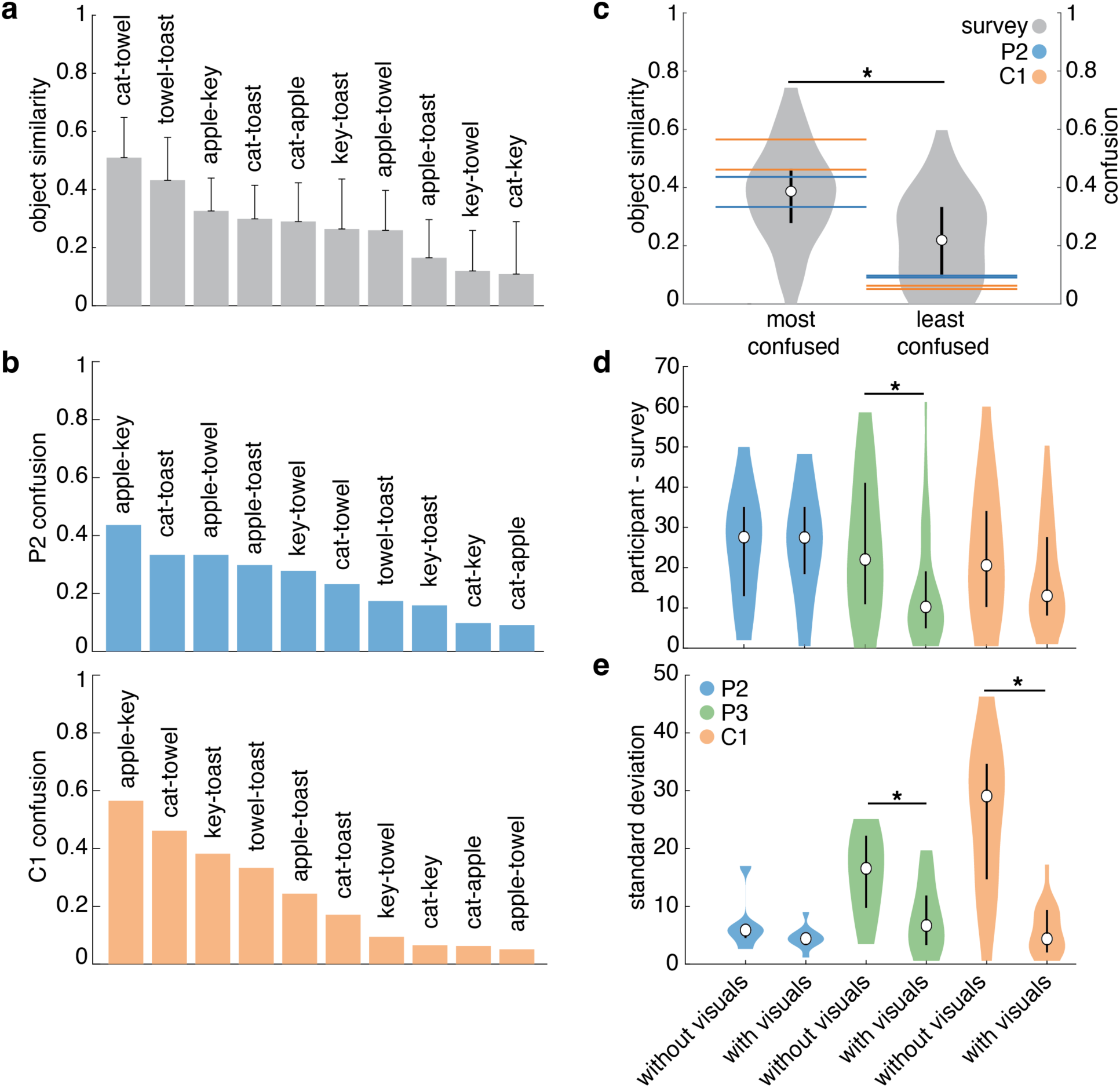
Expected and perceived object-specific tactile characteristics. **a.** Mean and standard deviation of object similarities between pairs of objects based on the survey reports of people with intact somatosensation. **b.** The fraction of times that each participant confused two objects with each other across all repetitions of a sensation. **c.** Horizontal lines show the participants’ confusion during the replay task for the two most confused objects (P2: apple-key and cat-toast, C1: apple-key and cat-towel) and the two least confused objects (P2: cat-apple and cat-key, C1: cat-apple and apple-towel). These lines correspond to the bars shown in b. The violin plots show the expected similarities (as shown in a.) between these objects based on the object quality survey results. The objects that participants P2 and C1 confused the most were rated by people with intact somatosensation as being significantly more similar to each other than the two objects that were confused the least. Participant P3 is excluded from this follow-up analysis due to their chance level performance on the replay task. **d.** Distribution of mean absolute differences between the survey ratings of participants P2, P3 and C1 with and without a visual context, and the survey ratings of the participants with intact sensation, across all objects. **e.** Distribution of mean standard deviations across all survey ratings of participant P2, P3 and C1 with and without an image that they thought best matched the evoked sensation. Median values are displayed as an open black circle and the 25-75% quartiles are displayed as a thick black line.

We wondered whether the visual presence of the object affected the verbal reports of the participants. To quantify this difference, the participants with spinal cord injury filled out the same survey (Fig. S5) as the participants with intact somatoensation in response to their created sensations with and without an image that they thought best matched the evoked sensation. We found mixed effects; only participant P3 significantly changed their ratings in the presence of visual information (p = 0.011, two-sample Kolmogorov-Smirnov test, α = 0.025, D = 0.40, Fig. 8d), whereas P2 (p = 1.000, D = 0.07) and C1 (p = 0.200, D = 0.27) did not. However, both P3 (p = 0.018, D = 0.49, Fig. 8e) and C1 (p = 0.002^-4^, D = 0.85, Fig. 8e) had more reliable survey responses when a visual context was provided. This was not true for participant P2 (p = 0.285, D = 0.47). These results suggest that when the stimulation-evoked sensations were not very distinct – P3 was no better than chance in the replay task and had large variations in selected stimulus parameters – the presence of a visual context skewed sensation reports towards the expected tactile characteristics based on the presented object image. In all other cases, as shown by the robustness of P2 and C1’s survey responses to a visual context and their significant replay performances, microstimulation of the somatosensory cortex alone can evoke distinct object-appropriate tactile perceptions.

## Discussion

In this study, people with microelectrode arrays implanted into area 1 of the somatosensory cortex were able to self-select complex microstimulation parameters to create sensations that mimicked tactile properties of natural objects. This was accomplished using a robust experimental paradigm that required participants to search through four stimulation parameters in a way that could not be learned, and that also provided a meaningful visual context for the five virtual objects (Fig. 1). While there was considerable overlap in the individual stimulus parameter sets for each object (Fig. 3), there were some significant differences for participants P2 and C1, which evoked perceptually distinct sensations that could be recognized in the absence of a visual context (Fig. 4a). The low performance of P3 is likely explained by insufficient parameter exploration in the object-sensation mapping task (Fig. 2c-f). The significant difference in exploration time between participants suggests that the parameters that P2 and C1 selected were indeed sought for their distinct sensations.

One could argue that participants simply tried to create distinct sensations – rather than intuitive or naturalistic ones – so that they could successfully perform the replay task. It would have been relatively easy for the participants to do this; the just-noticeable differences in amplitude and frequency are about 10-20 µA^22,23^ and 3-60 Hz^24^, respectively, and the amplitude and frequency steps they could control were 8 µA and 14 Hz, respectively. Creating five distinct sensations by manipulating one or two stimulation parameters should have been trivial. However, when the participants were asked to create sensations that represented each displayed object, the stimulation parameters could predict both compliance and temperature across the object set, reflecting the innate characteristics of these objects (Fig. 7). Further, objects with more similar tactile characteristics were more often confused with each other during the replay task (Fig. 8). These lines of evidence strongly suggest that the participants both attempted to and were successful at creating intuitive sensations; they did not seek to simply create distinct sensations. Moreover, the ability of an LDA classifier, trained on stimulation parameters alone, to predict object identity above chance, demonstrates that there was consistency in the parameter-selections across different days (Fig. 4a), and that participants P2 and C1 did not simply maximize a single parameter (Fig. 4b).

The stimulation parameters that the participants chose were highly variable within and across objects (Fig. 3). In contrast, the subjective tactile experiences of these objects, when paired with vision, were distinct and vivid (Fig. 5). This suggests that with the right context, ICMS-induced sensations can give rise to realistic touch experiences. Compared to previous research, this experiment was different in two key ways: participants were in control of their own stimulation and participants could actively explore an object that was presented visually. During passive stimulation (no vision, no exploration), people typically report skin-level sensations, such as “pressure” or “vibration”, as their attention is focused on their body^8,10^. During active exploration however, their attention is likely focused on the external world, thereby interpreting these same percepts as object-oriented sensations, such as compliance and roughness. The difference in experimental context could explain why the participants in this study spontaneously reported novel and more object-oriented sensation descriptors (Fig. 5). As people will ultimately use closed-loop brain-computer interfaces in daily life, it is important to investigate the functional use and experience of ICMS-evoked sensations in their relevant multi-sensory context.

### Limitations

Participants explored a large parameter space in a blinded fashion. Because the parameter space was randomized on each trial, participants could not learn a mapping between their movements in the space and their perceived sensory characteristics. Although this manipulation was crucial to ensure that participants made their choices based on tactile perception, rather than vision, it also made the experimental task very challenging, and may have contributed to the large variance in the stimulation parameters that represented particular objects. Although the object-specific stimulus trains were recognizable within a single session, participants could recognize only a few stimulus trains correctly that were selected from multiple previous sessions. As such, the significantly larger variation in stimulus parameters across compared to within sessions seemed to complicate replay performance (Fig. S3). However, only a limited number of stimulus trains were tested in these across-session replay tests and additional research is needed to better determine how the sensation quality may change across days.

The discrepancy between the participants’ certainty (Fig. 6c) and replay performance (Fig.4) may be due in part to the strictness of the replay task. It may be the case that participants were correct about some perceptual qualities and wrong about others. Participants often confused sensations for more similar objects (Fig. 8b) and performed better at recognizing the compliance and temperature differences between objects than recognizing the object itself (Fig. 7). Rather than getting some recognition for the correctly recognized qualities in the replay task, their object selection is simply counted as incorrect. Other forms of evaluation may be better suited to assess the experienced quality of the created sensations. For example: rating the similarity between the artificial sensation and an actual sensation on a sensate area of the hand or grouping sensations with similar qualities together irrespective of their (potentially incongruent) visual representation.

## Outlook

Brodmann’s area 1 is responsible for processing tactile input from mechanoreceptors that give rise to percepts like texture^11,25–27^. By activating this region of the brain using electrical stimulation, we have the ability to evoke intuitive sensations that may resemble features of naturally evoked touch^16^. This study shows that stimulation can evoke intuitive percepts that represent a variety of tactile object properties and that visual input contributes to the overall experience of artificial touch. In the future, self-guided stimulation approach may be used to efficiently characterize percepts evoked by novel stimulation paradigms.

## Online Methods

### Participants

This study was part of a multi-site clinical trial, registered at ClinicalTrials.gov (NCT01894802). The purpose of this trial is to collect preliminary safety information and demonstrate that intracortical electrode arrays can be used by people with tetraplegia to both control external devices and generate tactile percepts from the paralyzed limbs. This manuscript presents the analysis of data that were collected during the participants involvement in the trial but does not report clinical trial outcomes. The study was conducted under an Investigational Device Exemption from the U.S. Food and Drug Administration and ethically approved by the Institutional Review Boards at the University of Pittsburgh and the University of Chicago. Ten years prior to the time of implantation, participant P2 sustained a C5 motor/ C6 sensory ASIA B spinal cord injury. He was between 25-30 years old at the time of implant and between 30-35 years old during the data collection for this study. Participant P3 was of similar age as P2 during implant and data collection and sustained a C6 ASIA B spinal cord injury 12 years prior to implantation. Participant C1 sustained a C4 ASIA D spinal cord injury 35 year prior to implantation. He was 55-60 years old at the time of implant and during data collection. Prior to any study procedures, all three participants provided their informed consent.

### Stimulation apparatus

Prior to this study, participants were implanted with two 2.4 x 4 mm microelectrode arrays (Blackrock Microsystems, Salt Lake City, UT) in Brodmann area 1 in their left hemisphere^10,28,^. Another two arrays were implanted in their motor cortex within the same hemisphere, but were not used in this study. Each sensory array contained 60 1.5 mm long electrode shanks, wired in a checkerboard pattern across a 6 by 10 grid such that 32 electrodes could be stimulated. The electrode tips were coated with a sputtered iridium oxide film. The stimulation return electrode was a titanium pedestal that was fixed to the skull. Upon stimulation, participants reported sensations on their right hand. The quality and location of these sensations depends on the stimulation protocol^9,10,17^. The stability of these sensations is measured each month using a survey. To do so, each stimulation electrode is stimulated one-by-one with a standard stimulus (60 μA, 100 Hz, 1 second), after which the participant reports the experienced location and quality of the resulting sensation (if any). Based on these historical data, a single set of three electrodes was selected per participant for the current study. These electrodes were chosen to maximize the reliability and intensity of the evoked sensation, meaning that a clearly detectable sensation was evoked (almost) every time the electrode was stimulated. Furthermore, these electrodes were chosen to evoke sensations across the palmer side of the right hand (Fig. 1b).

A CereStim C96 multi-electrode microstimulation system (Blackrock Microsystems, Salt Lake City, UT) was used to deliver stimulation. Each stimulation pulse was current-controlled and charge-balanced, consisting of a 200 μs cathodal phase followed by a 100 μs interphase period and a 400 μs anodal phase set to half of the amplitude of the cathodal phase. Electrodes could be stimulated up to 300 Hz at amplitudes ranging from 2 to 100 μA^10,23,29^.

### Stimulation protocol

During this study, participants were able to manipulate four ICMS parameters in real-time: amplitude, frequency, biomimetic factor and drag (Fig. 1c). Each of these parameters could be manipulated across 10 levels. The minimum amplitude was set to 10 μA for all participants, which is just below or at detection threshold for most electrodes^17^. Predetermined individual safety limits set the maximum amplitude to 100 μA for P2, 80 μA for P3 and 90 μA for C1.

To accommodate for potential differences in the perceived intensity of a stimulus at the same stimulus current across electrodes, participants were asked to equalize their perceptual intensity across the three electrodes at the start of each session. One electrode would serve as a base with a fixed amplitude of 48 μA for P2 and 44 μA for P3 and C1. Participants were asked to match the perceived intensity of the other two electrodes to the base electrode using two sliders that manipulated their respective amplitudes. The participants could stimulate each electrode for 1s whenever they wanted to. Participants were not told that they were manipulating the amplitude level of different electrodes. Moreover, the electrode assignment to stimulation buttons 1, 2 or 3 was randomized so participants were not aware what electrode they were manipulating. Each electrode served as base electrode once during a block of three trials. Based on the resulting amplitude ratio, an individual amplitude range was determined for each electrode. For example, with an amplitude ratio of 48(base):40:60 = 0:-8:+12 μA, the ranges for each electrode would be: electrode 1 = [18, 88] μA, electrode 2 = [10, 80] μA, and electrode 3 = [30,100] μA. The final amplitude ranges were set to the mean across the amplitude ranges obtained at each of the three trials. For each electrode, 10 values were chosen that equally divided the determined amplitude range, e.g., electrode 1 = [18, 26, 34, 42, 50, 56, 64, 72, 80, 88] μA. To control for possible differences in the perceived intensity that may occur over the course of a single session, the task was repeated at the end of the session. No significant differences were found between the task results at the beginning and end of a session (Fig. S6a).

The stimulation frequency was divided into ten increments between 20 and 150 Hz and the same frequency was used on all three electrodes in all trials. Because frequency may have electrode-dependent effects on perception^17^, we delivered single-electrode stimulus trains at 20, 85 or 150 Hz and asked participants to indicate on a screen which stimulus was the most intense. This was done for all three electrodes in a random order at an amplitude of 62 μA and this task was repeated at the end of the session. Although participant C1 consistently rated a high frequency (150 Hz) stimulus as most intense on all electrodes, the results of participants P2 and P3 were mixed. Participant P2 seemed to have a low-frequency (20 Hz) preferring electrode, a high-frequency preferring one and a medium-frequency (85 Hz) preferring one. Participant P3 seemed to have two high-frequency preferring electrodes and one medium-frequency preferring one. Except for electrode 3 of participant P2, there was no significant difference between the task results at the start and end of a session, suggesting that the frequency and electrode specific differences in intensity were consistent over time (Fig. S6b).

The drag parameter determined how long an electrode remained active after the cursor left its receptive field on the digital object. The total time that an electrode was stimulated was calculated as: *E* = *T* + *D* ∗ *T*, where *T* was the total time that the cursor was on the receptive field of that electrode (Fig. 1b), and *D* was a participant-selected drag factor between 0 and 2. By increasing drag, multiple electrodes could be active at the same time.

Neural activity in the somatosensory cortex typically contains a clear onset and offset transient in response to a mechanical indentation of the skin^15^. The magnitude of the onset transient is about 15 times as big as that during the sustained period of skin contact, whereas the offset transient is about 8 times as big. We mimicked these naturally occurring patterns of neural firing activity via a biomimetic factor that specified the amplitude difference between the onset and offset transients and the amplitude during sustained object contact. The new amplitude was calculated as 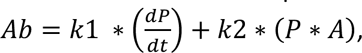 where *k*1 was defined as 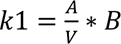 for the onset and 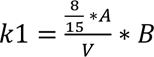 for the offset transient, *B* was the participant-selected biomimetic factor between 0 and 10, *dP* was the change in pixel color as the cursor moved across an object mask (Fig. S7), *dt* a fixed time interval of 20 ms, *k*2 determined the maximum amplitude during sustained touch as 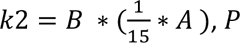 the current masked pixel color (Fig. S7), *A* the participant-selected amplitude, and *V* was a constant of 5 that indicated the maximum expected velocity. For each object, a black and white mask was created, defining an onset and offset gradient that was appropriate for its corresponding compliance level (Fig. S7). The pixel values were taken from this mask image. When the biomimetic factor was at its maximum, it was maximally mimicking the biologically inspired amplitude where the onset transient was set at a maximum of 15x the amplitude during sustained touch and the offset transient is at maximum 8x the amplitude during sustained touch. In case of an unexpected drastic velocity increase, the amplitude could never exceed the participant-selected amplitude level. The amplitude during sustained touch was set to 1/15x the participant-selected amplitude.

### Experimental procedure

Our experimental design is inspired on Shokur et al.^19^, who asked participants with paralysis of both legs due to spinal cord injury to create sensations for walking on grass, pavement or sand using vibro-tactile stimulation on their lower arms. Similar to Shokur et al., four different ICMS parameters were mapped to the x- and y-axes of two rectangles. Due to the limited movement capabilities of our participants, the rectangles were presented on a Windows Surface laptop (Fig. 1a). By moving around a cursor in each rectangle, participants could adjust the stimulation parameters in real time. Above the rectangles, one of five objects was displayed: apple, toast, towel, cat, or key (Fig. 1a,d). Participants were asked to find the best possible stimulation settings that represented these objects while they interacted with it; we called this the “object-sensation mapping task.” Participants could trigger ICMS by touching the virtual object using a stylus, or by using an automatic stimulation option that caused a digital cursor to move back and forth across the object every 6.5s, roughly evoking stimulation for 4s with a 2.5s break (Fig. 1a). Hidden from the participants view, the presented object was divided into three regions: as long as the cursor was within one field, a specific electrode would be stimulated (Fig. 1b).

For each participant, we selected three electrodes that evoked sensations in the fingers and or palm of the right hand^10,17^. Moving a cursor across the digitally presented object created a sensation across the palmer side of the right hand (Fig. 1b). Allowing participants to make their own explorative movements across the object was done to create a more realistic and interactive experimental setting. Depending on how participants moved the cursor across the presented object, they could influence the location, biomimetic factor, drag and overall length of stimulation. As our participants still had some control of their arms, they performed this task using the back of their hand or a stylus. To prevent actual sensory input from interfering with the ICMS-evoked sensations, participants controlled the interface using their left hand.

Each object was repeated two to three times per session. Each trial, the parameter assignment to the rectangle axes was randomized. In addition, each parameter distribution was randomly flipped (i.e., running from minimum to maximum, or from maximum to minimum on a particular rectangle axis). This randomization forced the participants to base their decisions on their sensation alone, rather than some memorized visual position of the cursors within the rectangles. At the end of each trial, after participants finalized a stimulation setting for an object, they were asked to rate how satisfied they were with their created sensation using a slider that ran from “unsatisfied” to “satisfied” (Fig. 1c). Across 22 sessions, participant P2 created 30 cat, key and toast sensations, 33 apple sensations and 29 towel sensations. Across 10 sessions, participant P3 created 20 cat and toast sensations, 18 apple and towel sensations, and 19 key sensations. Similarly, participant C1 created 21 cat, 19 apple, key and towel sensations and 20 toast sensations across 10 sessions.

To test whether the sensations were unique and distinguishable for the different objects, the stimulus parameters sets for each object in a session (regardless of their satisfaction score) were replayed to the participants at the end of that session. In this ‘replay task’, the participants experienced the ICMS-evoked sensations without any corresponding visual stimulus; a grey rectangle was shown instead of the original object (Fig. 1d). Participants were then asked to indicate what object best matched the experienced sensation. After each object selection, they were asked to indicate how certain they were of their choice, using a slider that ran from “uncertain” to “certain”. A total of two to three repetitions per unique sensation were collected per session.

To investigate the stability of the sensations across time, as well as their dependence on visual context, participants completed two additional sessions of the replay task three (P3, C1) to seventeen (P2) weeks after finishing data collection. During these sessions, participants were presented with three parameter sets per object: the sensation with the highest satisfaction rating in the first and second halves of the experimental sessions, and the sensation that was closest to the median of the selected parameters for that object. First, participants were asked to rate (Fig. S5) the experienced sensations using the same tactile dimensions as used in the online survey with control participants (Fig. S1a). Then, participants had a chance to familiarize themselves with the previously created sensations and go through each of them at their own pace while being presented with their correct visual context. Lastly, participants performed a modified version of the replay task in which they could select and explore the object image that they thought best matched the evoked sensation. After assigning an object to a sensation, thus providing their own visual context, participants were asked to fill out the same survey as before (Fig. S5). This replay task was repeated another two times without the survey, providing a total of three repetitions per unique sensation. Finally, during one of the sessions, all participants completed a short survey on their experience during the experiment (Fig. S4).

### Statistical analysis

A two-sided Kolmogorov-Smirnov test was used to test whether the collected pre- and post-session amplitude ranges differed significantly per participant. The significance level was set to 0.025, as a single comparison was made per participant. A similar strategy was used to evaluate the pre- and post-session frequency intensity choices per electrode. However, as these differences were assessed for each of the three stimulation electrodes per participant, the significance level was Bonferroni-corrected and set to 0.025/3 = 0.008.

We compared the trial completion times, aggregated stimulation times per trial and total percentage of explored parameter space across participants using two-sided Wilcoxon rank sum tests. A single test was performed for each of these three measured variables. After Bonferroni correction^30^, the significance level was set at 0.025/3 = 0.008.

To assess differences between stimulation parameters that were chosen during the object-sensation mapping task, we used a two-sided Kruskal-Wallis test to assess whether the individual parameter selections for different objects came from the same or a different distribution. We only included trials that had a normalized satisfaction score of at least 50 out of 100 were included in the analysis. To compensate for multiple comparisons across each of the four stimulation parameters, the Bonferroni corrected significance level was set to 0.025/4 = 0.006. If a significant difference was found, post-hoc one-sided two-sample Kolmogorov-Smirnov tests were used to determine what object-specific parameter selections were different from each other, testing each possible combination of two objects. The Bonferroni-corrected statistical significance level was set to 0.05/10 = 0.005.

To determine whether a specific object could be predicted from a specific stimulation parameter set, a linear discriminant analysis (LDA) classifier was trained and tested on the object-specific parameter selections of each participant using 10-fold cross validation. Because the sample size in this study was small, collecting only a maximum of 30 samples per object, the 10-fold cross validation procedure was repeated 1000 times and the results were averaged to get a stable approximation of the true classifier performance. Moreover, to compensate for unequal samples per object, each class sample was supplemented to match the maximum class size with randomly drawn samples from the same object distribution. This was repeated on each of the 1000 repetitions of the 10-fold cross-validation procedure. A permutation test with 1000 permutations was then used to assess a statistically significant classification performance^31^. For each permutation, a random sample of the maximum class size was drawn without replacement from all stimulation-parameter selections. Then, 10-fold cross validation was performed on this sample, generating a null distribution of possible LDA classification performances. The LDA performance was considered significant if it was greater than 95% of these classification performances. To check for the individual contributions of each stimulation parameter to the significant LDA performances, an LDA classifier was trained and tested on various combinations of stimulation parameters using the same procedure for amplitude, amplitude + frequency, amplitude + frequency + biomimetic factor, and amplitude + frequency + biomimetic factor + drag.

To check whether any differences in stimulation parameter selections could be explained by their relative contribution to the total charge per electrode, differences in total charge per object were calculated based on the replay data. Because participants were free to explore these sensations as they wished by touching a virtual rectangle, there were no fixed-length stimulation trains. To compensate for relative differences in total stimulation time between electrodes, the total charge values were normalized to the total time spent on each electrode. A two-sided Kruskal-Wallis test was used to assess any significant differences in the total charge between different objects. The statistical significance level was set to 0.025. In case a statistical difference was found, multiple post-hoc one-sided two-sample Kolmogorov-Smirnov tests were applied to assess what objects were significantly different from each other. The Bonferroni-corrected statistical significance level was set to 0.05/10 = 0.005.

The participants’ performance on the replay task was assessed by calculating the average percentage of correct answers (the chosen object equals the original object for which the sensation was designed) across all sessions. To assess significance, the participant’s performance was compared against a naïve classifier that predicted each possible class with equal probability. Using a permutation test with 1000 permutations, the performance of this naïve classifier was calculated against the underlying true class distribution across all trials. Chance level was established as the mean performance of this classifier across all permutations. The participant’s task performance was considered significant if the percentage of correct answers across all trials exceeded 95% of all permuted naïve classifier performances. A one-sided nonparametric Wilcoxon rank sum test was used to assess whether the participants certainty scores were significantly higher in case their answers were correct compared to incorrect^32^. The significance level was set to 0.05, as a single comparison was made per participant.

The participants’ performance on the across session replay task of the final two sessions was assessed in a similar way to the within session replay performance. In addition, we calculated which sensations were correctly matched to their original target in at least two out of three repetitions. To assess the variation of object-specific stimulus parameters within and across sessions, we calculated the standard deviation across all repetitions of an object-specific parameter within and across sessions for each participant. A one-sided Kruskal-Wallis test (with a significance level of 0.05) was used to assess whether the object-specific variation across all parameters and participants was significantly different within and across sessions. Additional post-hoc one-sided two-sample Kolmogorov-Smirnov tests (Bonferroni corrected significance level was set to 0.01) were used to assess significant differences in the variation of each individual parameter within and across sessions.

Satisfaction scores were normalized to the minimum and maximum scores per participant to account for individual differences. A two-sided nonparametric Kruskal-Wallis test was used to assess whether the satisfaction ratings across all participants came from the same or a different distribution per object^33^. If a statistically significant difference was found, individual post-hoc one-sided nonparametric two-sample Kolmogorov-Smirnov tests were used to assess which objects differed significantly from each other in terms of their satisfaction rating^34^. To compensate for multiple comparisons, testing potential differences between all 10 possible combinations of five object sensations, Bonferroni correction was used^30^. After Bonferroni correction, the level of significance was set to 0.05/10 = 0.005.

Another classification strategy was used to assess whether tactile characteristics, rather than objects, could be distinguished based on the stimulation parameter selections. To do so, five individual binary LDA classifiers were trained and tested to predict different levels of compliance, friction, temperature, micro- and macrostructure. Each tactile characteristic was divided into two levels: minimum (e.g., “smooth”) and maximum (e.g., “rough”). These levels were determined by the mean ratings on these tactile dimensions being bigger or smaller than 0, as judged by the participants with intact somatosensation (Fig. S1). Each individual classifier was trained and tested using the same 10-fold cross validation procedure and permutation test as described above. For each object, the percentage of classifier predictions of each tactile characteristic was computed.

To assess the influence of visual context on the experience of the customized ICMS-evoked sensations, we calculated the absolute difference in survey ratings between our participants and the median of the survey results from people with intact somatosensation for each object. We did this for the sensations that we presented with and without a self-selected visual context in the final two replay sessions of our participants. In addition, we calculated the standard deviation across the survey ratings of our participants for each object-specific sensation. Significant differences in the absolute mean difference between our participants’ and the control participants’ ratings, and in the standard deviation in survey ratings across repeated presentations of object-specific sensations were assessed using a two-sided two-sample Kolmogorov-Smirnov test. Significance level was set to 0.025 as a single test was performed on each dataset.

Lastly, the Euclidean distance of the control participants’ survey ratings was calculated for each possible pair of objects. These distance measurements were then normalized to the maximum object-pair distance. To get a measurement of object similarity rather than difference, we subtracted these normalized Euclidean distances from 1 for each object pair. This provides a similarity score between 1 (most similar) and 0 (most dissimilar). Next, we calculated the percentage of object-specific sensations that was wrongfully assigned to the other object for each possible object pair. For example, in case of apple and cat sensations, we calculated the percentage of apple sensations that were wrongfully assigned to a cat object plus the percentage of cat sensations that were wrongfully assigned to an apple across all presentations of apple and cat sensations during the regular replay task. We then compared the control participants’ similarity scores corresponding to the two most and least confused object pairs of each participant. A two-sided two-sample Kolmogorov-Smirnov test was used to assess whether the most confused objects corresponded to the most similar ones. Significance level was set to 0.025 as a single test was performed.

## Data Availability

All data produced in the present study are available upon reasonable request to the authors.

## Acknowledgements

We would like to thank the magnificent N. Copeland, Mr. Dom, and S. Imbrie for their participation in this research study. Without these wonderful brain-computer interfacing pioneers, this research would not be possible. In addition, we would like to acknowledge Prof. Bensmaia’s support of this project; his critical thinking and team spirit that have been crucial to this project’s success. This study was supported by the National Institute for Neurological Disorders and Stroke (UH3 NS107714) and the Dutch Research Council (NWO Rubicon: 019.193SG.011, NWO Vidi: VI.Vidi.191.210).

## Author contributions

Under the supervision of BS and RG, CV led the experiment design, implementation, data collection and analysis, and wrote the first draft of the research article. VK conducted part of the experiment implementation and data collection. CG conducted the data collection with participant C1. SB and MB provided feedback on the data analysis. All authors provided critical review of the text.

## References

1. Bensmaia, S. J. & Miller, L. E. Restoring sensorimotor function through intracortical interfaces: Progress and looming challenges. Nat. Rev. Neurosci. 15, 313–325 (2014).

2. Nghiem, B. T. et al. Providing a Sense of Touch to Prosthetic Hands. Plast. Reconstr. Surg. 135, 1652–1663 (2015).

3. Perruchoud, D., Pisotta, I., Carda, S., Murray, M. M. & Ionta, S. Biomimetic rehabilitation engineering: The importance of somatosensory feedback for brain-machine interfaces. J. Neural Eng. 13, (2016).

4. Collinger, J. L., Gaunt, R. A. & Schwartz, A. B. Progress towards restoring upper limb movement and sensation through intracortical brain-computer interfaces. Curr. Opin. Biomed. Eng. 8, 84–92 (2018).

5. Schiefer, M. A., Graczyk, E. L., Sidik, S. M., Tan, D. W. & Tyler, D. J. Artificial tactile and proprioceptive feedback improves performance and confidence on object identification tasks. PLoS ONE 13, e0207659 (2018).

6. Cordella, F. et al. Literature review on needs of upper limb prosthesis users. Front. Neurosci. 10, 209 (2016).

7. Pylatiuk, C., Schulz, S. & Döderlein, L. Results of an internet survey of myoelectric prosthetic hand users. Prosthet. Orthot. Int. 31, 362–370 (2007).

8. Armenta Salas, M., et al. Proprioceptive and cutaneous sensations in humans elicited by intracortical microstimulation. eLife 7, e32904 (2018).

9. Flesher, S. N. et al. A brain-computer interface that evokes tactile sensations improves robotic arm control. Science 372, 831–836 (2021).

10. Flesher, S. N. et al. Intracortical microstimulation of human somatosensory cortex. Sci. Transl. Med. 8, (2016).

11. Darie, R., Powell, M. & Borton, D. Delivering the Sense of Touch to the Human Brain. Neuron 93, 728–730 (2017).

12. Callier, T., Brantly, N. W., Caravelli, A. & Bensmaia, S. J. The frequency of cortical microstimulation shapes artificial touch. PNAS 117, 1191–1200 (2020).

13. O’Doherty, J. E. et al. Active tactile exploration using a brain-machine-brain interface. Nature 479, 228–231 (2011).

14. Zaaimi, B., Ruiz-Torres, R., Solla, S. A. & Miller, L. E. Multi-electrode stimulation in somatosensory cortex increases probability of detection. J. Neural Eng. 10, (2013).

15. Callier, T., Suresh, A. K. & Bensmaia, S. J. Neural Coding of Contact Events in Somatosensory Cortex. Cereb. Cortex 29, 4613–4627 (2019).

16. Lee, B. et al. Engineering artificial somatosensation through cortical stimulation in humans. Front. Syst. Neurosci. 12, (2018).

17. Hughes, C. L. et al. Perception of microstimulation frequency in human somatosensory cortex. eLife 10, (2021).

18. Hughes, C. L., Flesher, S. N. & Gaunt, R. A. Effects of stimulus pulse rate on somatosensory adaptation in the human cortex. Brain Stimulat. 15, 987–995 (2022).

19. Shokur, S. et al. Assimilation of virtual legs and perception of floor texture by complete paraplegic patients receiving artificial tactile feedback. Sci. Rep. 6, (2016).

20. Lee, S. D., Jung, Y., Chung, Y. A. & Lee, W. Neural substrates in secondary somatosensory area for the perception of different tactile sensations. Int. J. Imaging Syst. Technol. 26, 85–91 (2016).

21. Kim, L. H., McLeod, R. S. & Kiss, Z. H. T. A new psychometric questionnaire for reporting of somatosensory percepts. J. Neural Eng. 15, (2018).

22. Greenspon, C. M., et al. Biomimetic multi-channel microstimulation of somatosensory cortex conveys high resolution force feedback for bionic hands. bioRxiv 02, (2023).

23. Hughes, C. L. et al. Neural stimulation and recording performance in human somatosensory cortex over 1500 days. J. Neural Eng. 18, (2021).

24. Callier, T., Brantly, N. W., Caravelli, A. & Bensmaia, S. J. The frequency of cortical microstimulation shapes artificial touch. Proc. Natl. Acad. Sci. 117, 1191–1200 (2020).

25. Eck, J., Kaas, A. L. & Goebel, R. Crossmodal interactions of haptic and visual texture information in early sensory cortex. NeuroImage 75, 123–135 (2013).

26. Salinas, E., Hernández, A., Zainos, A. & Romo, R. Periodicity and Firing Rate As Candidate Neural Codes for the Frequency of Vibrotactile Stimuli. J. Neurosci. 20, 5503–5515 (2000).

27. Lieber, J. D. & Bensmaia, S. J. High-dimensional representation of texture in somatosensory cortex of primates. Proc. Natl. Acad. Sci. U. S. A. 116, 3268–3277 (2019).

28. Greenspon, C. M., et al. Tessellation of artificial touch via microstimulation of human somatosensory cortex. bioRxiv (2023).

29. Cogan, S. F. Neural stimulation and recording electrodes. Annu. Rev. Biomed. Eng. 10, 275–309 (2008).

30. Napierala, M. A. What is the Bonferroni correction? AAOS Now 40–41 (2012).

31. Ojala, M. & Garriga, G. C. Permutation Tests for Studying Classifier Performance Markus Ojala. J. Mach. Learn. Res. 11, 1833–1863 (2010).

32. Gibbons, J. D. & Chakraborti, S. Linear rank tests for the location problem. in Nonparametric Statistical Inference 298–307 (Marcel Dekker Inc., New York, 2003).

33. Ostertagova, E., Ostertag, O. & Kováč, J. Methodology and application of the Kruskal-Wallis test. In Applied Mechanics and Materials vol. 611 115–120 (Trans Tech Publications Ltd., 2014).

34. Berger, V. W. & Zhou, Y. Kolmogorov–Smirnov Test: Overview. in Wiley StatsRef: Statistics Reference Online (eds. Balakrishnan, N., et al.) (2014).

