## Supplemental Figure 1 for "Conveying tactile object characteristics through customized intracortical microstimulation of the human somatosensory cortex"

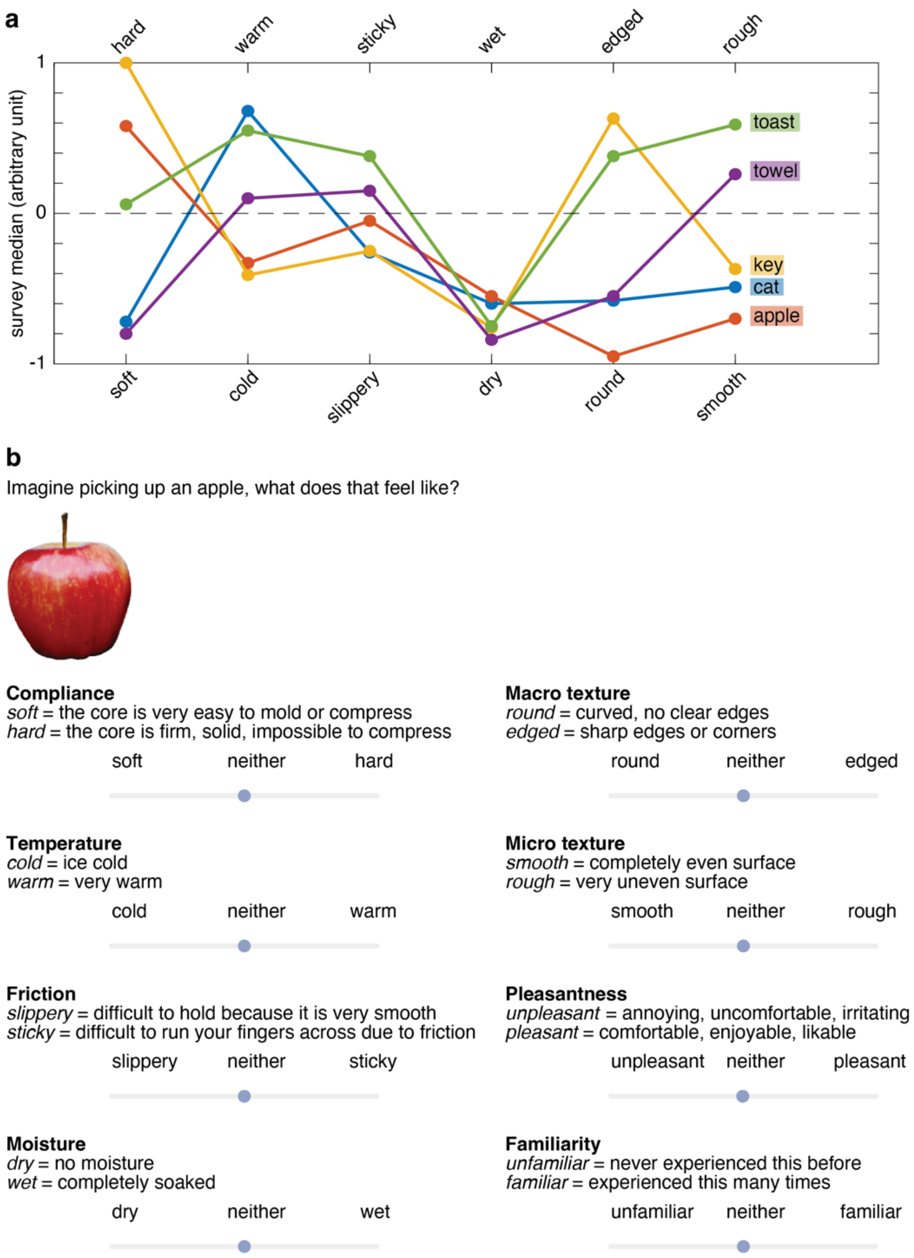


***Figure S1.*** *Tactile quality survey results of 34 people with intact somatosensation.* ***a.*** *Median levels of compliance, temperature, friction, moisture, micro and macro structure ascribed to each object according during the object quality survey.* ***b.*** *Example screenshot of a single item in the online survey. A total of 30 objects were presented, including an apple, banana peel, book, cat, cinnamon roll, glass, pair of glasses, pair of gloves, hammer, hand, ice cube, electric toothbrush, rock, key, knife, needle, orange, pencil, smartphone, rabbit, boiled rice, pair of scissors, pair of socks, used sponge, sweater, teddy bear, piece of toast, roll of toilet paper, towel and a wig. Each object was associated with a specific action: pick up, pet, shake, or grab. Participants were asked to rate the compliance, temperature, friction, moisture, macro texture, micro texture, pleasantness, and familiarity of these imagined object interactions.*
