## Supplemental Figure 2 for "Conveying tactile object characteristics through customized intracortical microstimulation of the human somatosensory cortex"

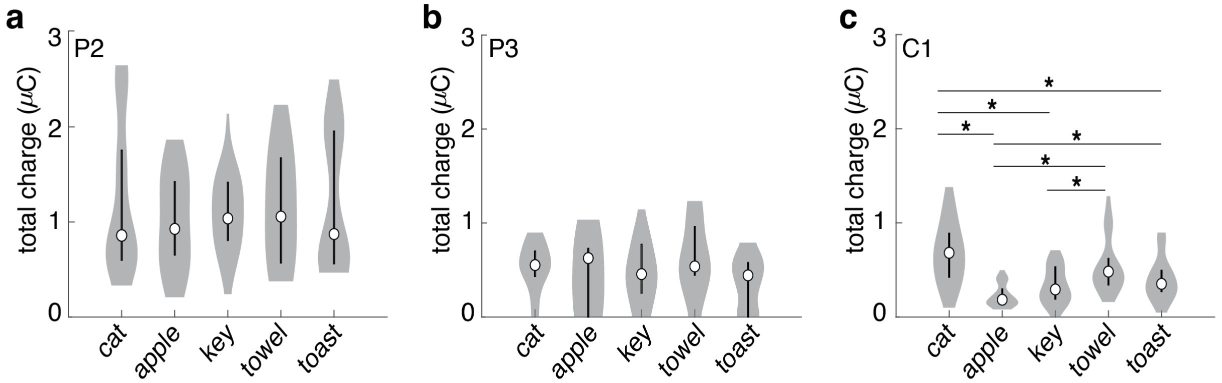


***Figure S2.*** *Normalized total charge of the object-specific stimulus for participant* ***a.*** *P2,* ***b.*** *P3 and* ***c.*** *C1. No significant differences in mean charge across all electrodes were found between the object-specific stimulation trains of P2 ($p = 0.768$, two-sided Kruskal-Wallis test, $\alpha = 0.025$, $\chi^2(4) = 1.83$) and P3 ($p = 0.238$, $\chi^2(4) = 5.52$). However, significant differences were found in the mean charge between the object-specific stimulation trains created by C1 ($p = 0.004^{-8}$, $\chi2(4) = 54.77$). C1 created higher charge (one-sided Kolmogorov-Smirnov test, Bonferroni corrected at $\alpha = 0.005$) stimulation trains for a cat compared to that of an apple ($p = 0.005^{-6}$, $D = 0.72$), key ($p = 0.001^{-2}$, $D = 0.56$) or toast ($p = 0.005^{-2}$, $D = 0.46$). Similarly, the charge created for a towel was higher than that of an apple ($p = 0.007^{-4}$, $D = 0.63$) and key ($p = 0.003$, $D = 0.41$), and the charge created for toast was bigger than that for an apple ($p = 0.002^{-1}$, $D = 0.48$). To check whether the total charge per electrode could explain the significant classification performances of P2 and C1, an additional LDA classifier was trained using 10-fold cross validation based on the total charge per electrode alone. This LDA classifier reached a performance of 20% ($p = 0.511$, permutation test, 1000 permutations without replacement, $\alpha = 0.05$) for P2, 18% ($p = 0.743$) for P3 and 34% ($p = 0.001$) for C1. These significant differences in total charge maybe due to C1's preferred use of automatic stimulation during the object-sensation mapping task. Whereas P2 and P3 used almost exclusively automatic stimulation, evoking an identical cursor movement across each object, C1 manually explored the objects to evoke stimulation. Moreover, C1 explicitly reported using different manual exploration strategies for each object, e.g., stroking a cat along it's back and drawing fast circles across a towel. The differences in cursor velocity per object will influence the result of the biomimetic factor differently, potentially enhancing differences in total charge between objects.*
