## Supplemental Figure 3 for "Conveying tactile object characteristics through customized intracortical microstimulation of the human somatosensory cortex"

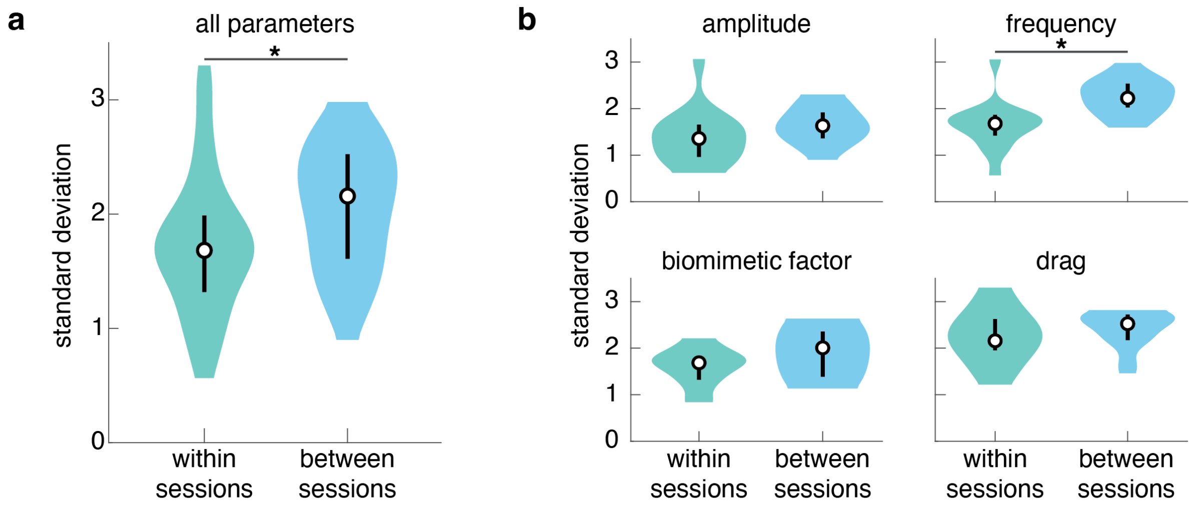


***Figure S3.*** *Distribution of the mean standard deviation of all within or across session repetitions of object-specific parameter selections.* ***a.*** *The distribution of object-specific standard deviations across all parameters and participants was significantly lower within sessions than across sessions.* ***b.*** *Same as in a, but shown for each individual stimulus parameter. The significant difference in within and across session parameter variation was explained by a significantly larger variation in the frequency parameter across sessions compared to within sessions ($p = 0.002^{-1}$, one-sided Kolmogorov-Smirnov test, Bonferroni corrected at $\alpha = 0.01$, $D = 0.73$).*
