## Supplemental Figure 4 for "Conveying tactile object characteristics through customized intracortical microstimulation of the human somatosensory cortex"

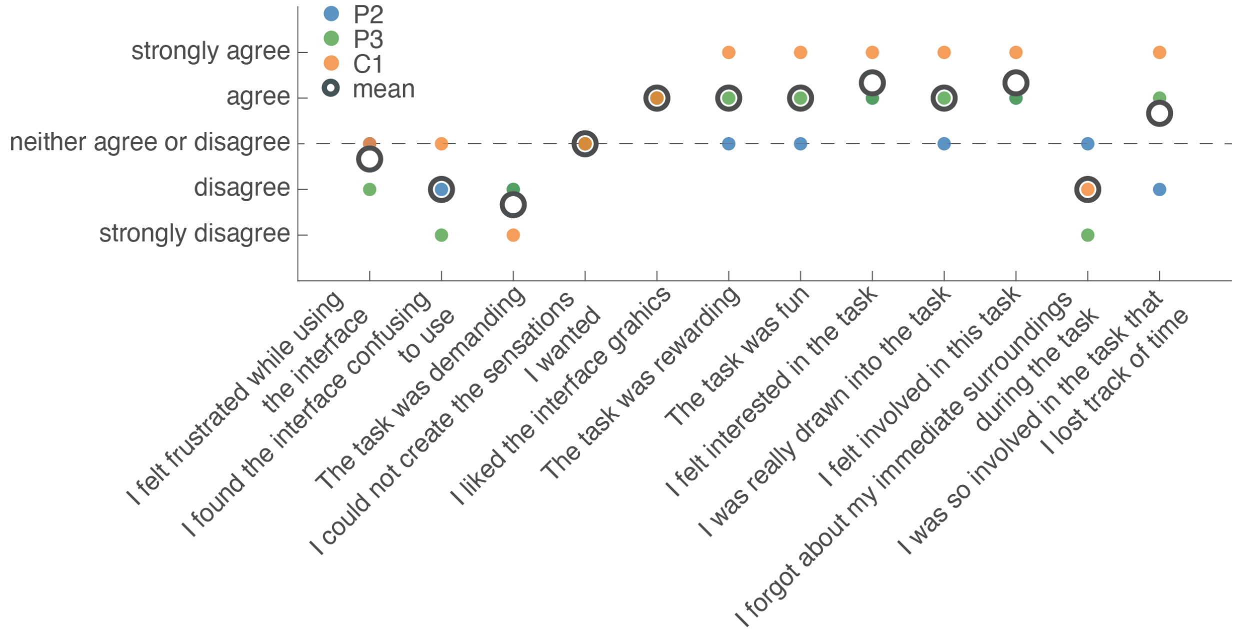


***Figure S4.*** *Participant engagement survey. The response of each participant is represented by a colored dot. The mean response across participants is highlighted by a black circle. Overall, participants were positive about their experiences using the tablet interface.*
