## Supplemental Figure 5 for "Conveying tactile object characteristics through customized intracortical microstimulation of the human somatosensory cortex"

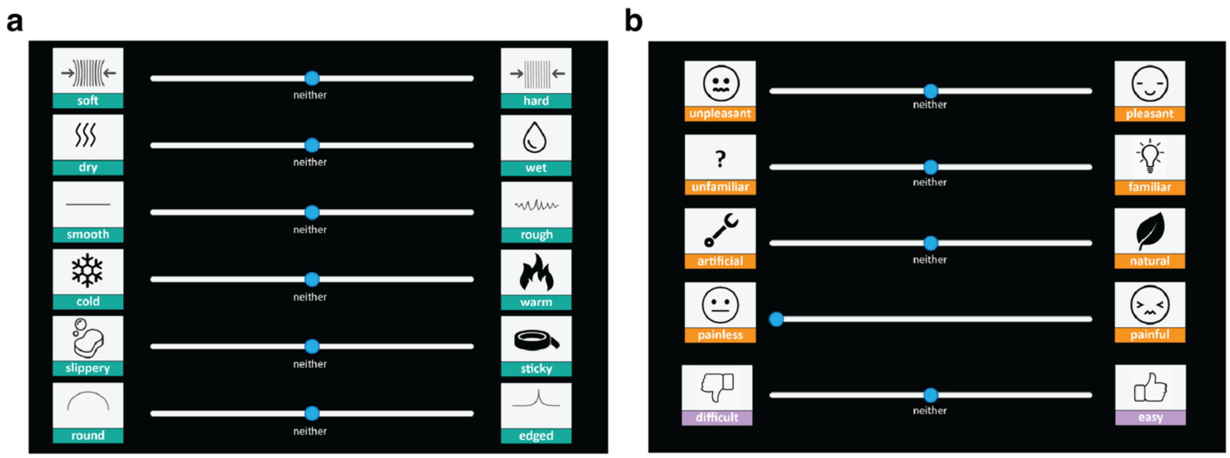


***Figure S5.*** *Screenshot of the tablet interface with the* ***a.*** *tactile and* ***b.*** *affective surveys. Participants could 'flip' the icon images to reveal a more elaborative description (identical to those used in Fig. S1) of the indicated tactile characteristic. Each slider (with the exception of the 'pain' one) started from a default position in the middle, indicating that this tactile characteristic was not experienced. Participants could slide it to the left or right to report their experienced sensations.*
