## Supplemental Figure 6 for "Conveying tactile object characteristics through customized intracortical microstimulation of the human somatosensory cortex"

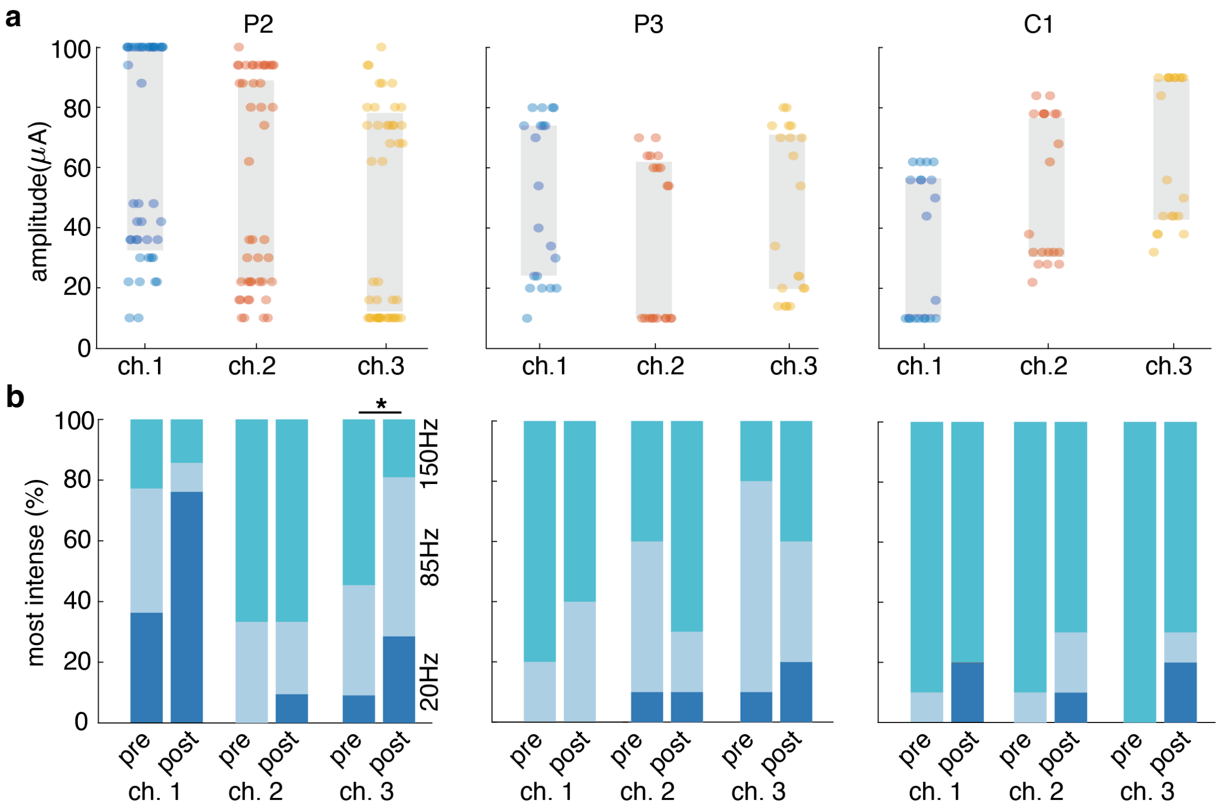


***Figure S6.*** *Pre-test parameter selections.* ***a.*** *Range of amplitudes used across all experimental test sessions per participant for the object-sensation mapping task. The grey rectangles indicate the mean minimum and maximum amplitudes per electrode. Clear differences were observed in the perceived intensity across the three stimulation electrodes. At the start of a session, participant P2 matched the electrode intensities with an average difference in amplitude of +11±10 μA between electrodes 1 and 2, and +20±13 μA between electrodes 1 and 3, P3 with a difference of +14±9 μA between electrodes 1 and 2, and +4±1 3μA between electrodes 1 and 3, and C1 with a difference of -20±4 μA between electrodes 1 and 2 and -32±6 μA between electrodes 1 and 3. We checked whether the results of this intensity matching procedure were consistent throughout a session and found no significant differences in the selected amplitude ratios between the start and end of a session (Kolmogorov-Smirnov test with P2: ch. 1-2 p = 0.423, ch. 1-3 p = 0.772; P3: ch. 1-2 p = 0.975, ch. 1-3 p = 1; C1: ch. 1-2 p = 0.675, ch. 1-3 p = 0.313).* ***b.*** *Total number of times that participants rated a low (20 Hz), medium (85 Hz) or high (100 Hz) stimulus frequency as most intense on a specific electrode. There was considerable variation across the intensity reports for all participants, e.g., although C1 rated a 150 Hz stimulus to be most intense on 80-85% of all trials on each electrode, he still selected a 20 Hz stimulus as most intense for 5-10% of the trials, and an 85 Hz stimulus as most intense on 5-15% of the trials. Except for electrode 3 of participant P2, the frequency and electrode specific differences in intensity were consistent at the start and end of a session (Wilcoxon signed rank test with P2: ch. 1 p = 0.025, ch. 2 p = 0.843, ch. 3 = .013*; P3: ch. 1 p = 0.366, ch. 2 p = 0.269, ch. 3 p = 0.705; C1: ch. 1 p = 0.503, ch. 2 p = 0.278 ch. 3 p = 0.078).*
