## Supplemental Figure 7 for "Conveying tactile object characteristics through customized intracortical microstimulation of the human somatosensory cortex"

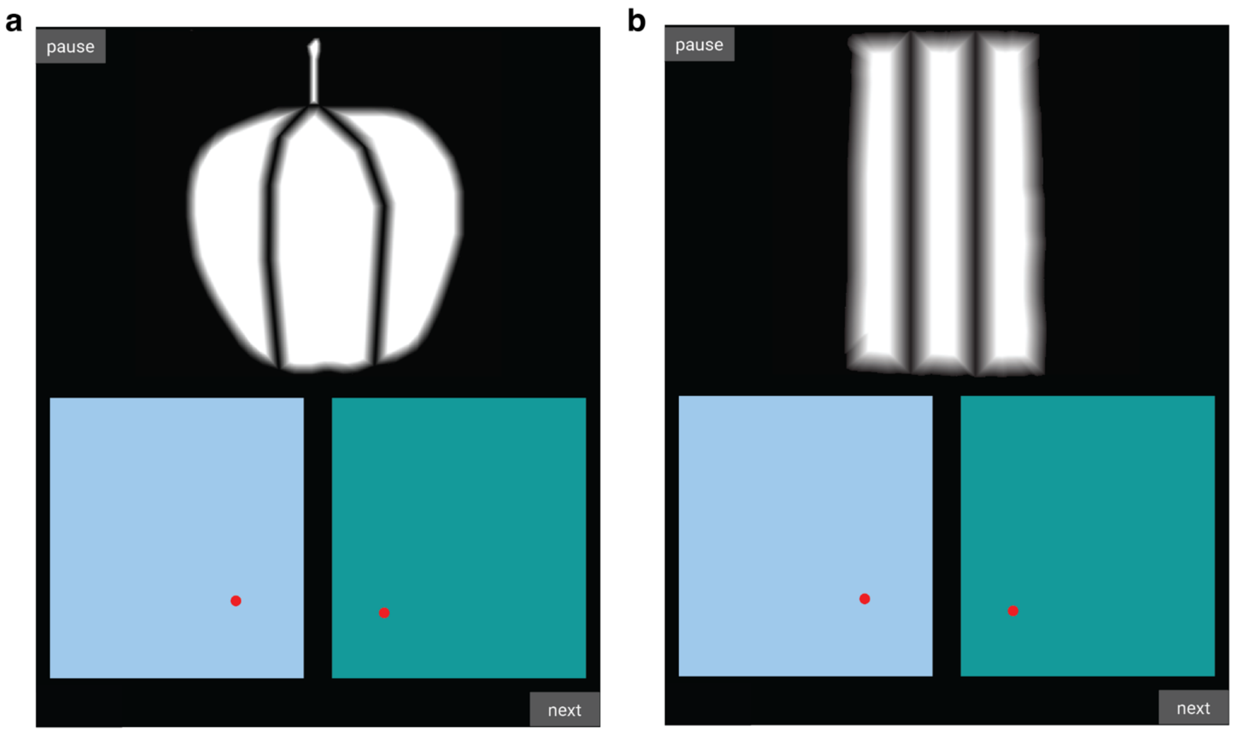


***Figure S7***. *Object masks for* ***a****. a hard (apple) and* ***b****. a soft (towel) object. The gradients created a pixel value for the on- and offset of each of the three electrodes "receptive field" on the displayed object. These object masks were used during the object-sensation mapping task. During the replay task, where the object was represented as a grey rectangle, identical gradients were used, albeit projected onto a rectangle rather than the original image shape.*
