## Supplemental Table 1 for "Conveying tactile object characteristics through customized intracortical microstimulation of the human somatosensory cortex"

**Table S1. Kruskal-Wallis test results of differences between individual stimulus parameters across objects. After Bonferroni correction, the significance level was set to 0.006. Asterisks denote statistically significant results.**

| Index | Participant | Variable | p-value | Test statistic |
|---|---|---|---|---|
| 1 | P2 | amplitude | $.001^{-1}$* | $\chi^2(4) = 23.16$ |
| 2 | P2 | frequency | .103 | $\chi^2(4) = 7.72$ |
| 3 | P2 | biomimetic factor | .003* | $\chi^2(4) = 16.13$ |
| 4 | P2 | drag | .004* | $\chi^2(4) = 15.5$ |
| 5 | P3 | amplitude | .458 | $\chi^2(4) = 3.63$ |
| 6 | P3 | frequency | .534 | $\chi^2(4) = 3.14$ |
| 7 | P3 | biomimetic factor | .296 | $\chi^2(4) = 4.92$ |
| 8 | P3 | drag | .137 | $\chi^2(4) = 6.98$ |
| 9 | C1 | amplitude | $.002^{-1}$* | $\chi^2(4) = 21.86$ |
| 10 | C1 | frequency | .063 | $\chi^2(4) = 8.92$ |
| 11 | C1 | biomimetic factor | .033 | $\chi^2(4) = 10.48$ |
| 12 | C1 | drag | .295 | $\chi^2(4) = 4.93$ |
