## Supplemental Table 2 for "Conveying tactile object characteristics through customized intracortical microstimulation of the human somatosensory cortex"

**Table S2. Permutation test results (1000 permutations without replacement) of tactile quality specific individual classifiers. The significance level was set to 0.05. Asterisks denote statistically significant results.**

| Index | Participant | Variable | p-value |
|---|---|---|---|
| 1 | P2 | compliance (63% accurate) | .009* |
| 2 | P2 | temperature (72% accurate) | .000* |
| 3 | P2 | friction (55% accurate) | .184 |
| 4 | P2 | micro structure (45% accurate) | .835 |
| 5 | P2 | macro struct. (53% accurate) | .355 |
| 6 | C1 | compliance (71% accurate) | .001* |
| 7 | C1 | temperature (76% accurate) | .000* |
| 8 | C1 | friction (63% accurate) | .072 |
| 9 | C1 | micro struct. (51% accurate) | .526 |
| 10 | C1 | macro struct. (47% accurate) | .777 |
